# Mapping sex differences in brain and cognition in relation to *APOE4* and amyloid burden: A longitudinal normative modelling study

**DOI:** 10.1101/2025.06.22.25330074

**Authors:** Sivaniya Subramaniapillai, Serena Verdi, Sarah Keuss, Kirsty Lu, Sarah-Naomi James, William Coath, David M. Cash, Frederik Barkhof, Marcus Richards, Andre Marquand, Jon Schott, James H. Cole, Ann-Marie de Lange

## Abstract

Sex differences in Alzheimer’s disease (AD) risk and progression are increasingly recognized, with females exhibiting higher global prevalence rates. Yet it remains unclear how genetic and biomarker indicators of Alzheimer’s risk, such as the apolipoprotein E-ɛ4 (*APOE4*) allele and amyloid burden, relate to sex differences in brain and cognitive health during the preclinical stage. Using established normative models trained on ~58,000 healthy participants, we computed regional z-scores from T1-weighted MRI scans in 372 cognitively normal participants from the Insight 46 cohort. Scans were acquired at two timepoints, approximately three years apart, beginning at age 70. Regions with z-scores below −1.96 were classified as brain-structure outliers and summarized as total outlier count (tOC). We used linear mixed-effects models to examine how sex, age, and AD risk (*APOE4* status and amyloid burden) relate to tOC and cognitive outcomes measured by Preclinical Alzheimer Cognitive Composite (PACC) scores. Both cross-sectional associations and longitudinal changes in tOC and PACC scores were examined, and we tested whether the effects of *APOE4* status and amyloid burden on brain and cognitive measures differed by sex. Cross-sectional analyses showed that males had greater tOC than females at younger ages. At timepoint 1, spatial maps showed more regions with outliers in males, though high outlier proportions were limited to occipital areas. By timepoint 2, group differences became more spatially distinct, with males and females showing deviations in different regions. Longitudinally, older males exhibited steeper increases in tOC over time compared to females. Females showed higher PACC scores overall, while no sex differences were observed in cognitive change over time. Greater tOC and amyloid burden were both associated with poorer cognitive outcomes, with the strongest association observed in female *APOE4* carriers. However, we found no evidence that AD risk influenced age-related changes in tOC or cognition over time. These findings highlight the complex interplay between sex, age, and AD risk in shaping brain structure and cognition in later life. Some of the observed patterns may reflect emerging vulnerability not yet captured by short-term longitudinal change, underscoring the importance of continued observation. In conclusion, normative modelling provides a valuable approach for detecting subtle variation in brain and cognitive aging across risk groups.

## Introduction

Sex differences in Alzheimer’s disease (AD) risk and prevalence are well-documented (Castro-Aldrete et al., 2025; Mielke, 2018; Niu et al., 2017; O’Neal, 2024). Females experience higher mortality rates due to AD worldwide (Huque et al., 2023; Nichols et al., 2019), though some countries and regions show higher rates among males (GBD Collaborators, 2019; Piovezan et al., 2020). Risk factors for AD, such as the apolipoprotein E-*ɛ*4 *(APOE4)* genetic variant, influence brain health differently across sexes (Brady et al., 2024). For example, females with the *APOE4* allele are at greater risk of developing AD at younger ages (Neu et al., 2017) and exhibit higher levels of amyloid β and tau pathology (Nemes et al., 2023; Sundermann et al., 2020). Understanding how genetic risk and early disease markers affect brain health trajectories in males and females during the preclinical phase is critical, as AD related brain changes often emerge long before the onset of overt clinical symptoms (Younes et al., 2019).

Amyloid-β accumulation is linked to an increased likelihood of developing AD, particularly when tau pathology is also present (Dang et al., 2018; Mormino & Papp, 2018; van der Kall et al., 2021). In amyloid PET studies of clinically normal older adults, findings show higher retention in females (Luchsinger et al., 2020; Palta et al., 2021; Sundermann et al., 2018), or no significant sex difference in global neocortical retention (Buckley et al., 2019; Rowe et al., 2010). Tau-PET imaging more consistently shows greater tau retention in females across middle-aged and older adults (Coughlan et al., 2025; Luchsinger et al., 2020; Palta et al., 2021; Wisch et al., 2021), with some studies indicating higher entorhinal tau retention in females with elevated amyloid levels (Buckley et al., 2019) or greater tau retention independent of amyloid (Digma et al., 2020).

AD pathology is linked to cognitive deficits, as measured by tools like the Preclinical Alzheimer Cognitive Composite (PACC), a measure for detecting cognitive decline in preclinical AD (Klinger et al., 2024; Mormino et al., 2017). As women tend to perform better on cognitive tests commonly used to assess early AD, particularly those assessing verbal memory and processing speed, early signs of pathology might be masked in women, raising concerns about the sensitivity of such assessments to sex-specific trajectories of decline (Sundermann et al., 2017). For example, females with mild cognitive impairment (MCI) have demonstrated better cognitive performance or comparable clinical symptoms than men, despite having greater AD pathology (Digma et al., 2020; Edwards et al., 2021; Sundermann et al., 2019). Notably, a longitudinal study in a North American cohort of clinically normal older adults found that females with higher amyloid-beta levels experience faster cognitive decline than males with comparable amyloid pathology (Buckley et al., 2018), highlighting the need for more studies to understand sex differences in long-term trajectories of brain and cognitive health.

Prior research on sex differences in AD has largely relied on cross-sectional cohorts from North America, often concentrating on specific brain regions, such as the temporal lobe, which is implicated early in AD. However, the significant heterogeneity in AD brain pathology, including contributions from vascular factors and other age-related processes, highlights the need to characterize structural changes both regionally and globally using metrics that capture individual differences that may be obscured in group-average maps (Habes et al., 2020; Verdi et al., 2023). Moreover, focusing on the preclinical stage, when participants are still cognitively normal, could provide an opportune window for early interventions that could delay disease progression. Normative modelling offers a robust framework for characterizing how brain regions are expected to appear across the healthy aging spectrum based on variables such as age and sex (Verdi et al., 2023). By training a model to predict specific brain features, such as cortical thickness or brain volume, we can identify individuals who deviate, either positively or negatively, from these normative expectations. In addition to identifying specific brain regions that deviate from the norm, the total negative outlier count (tOC) provides a global measure of the cumulative number of brain regions exhibiting negative deviation. While normative models have been applied to dementia patients, including AD (Loreto et al., 2024; Verdi et al., 2023, 2024) and Lewy body dementias (Bhome et al., 2024), to our knowledge, there has been no explicit exploration of sex differences and the role of AD risk factors in brain aging among cognitively normal individuals in a population-based longitudinal cohort.

In this study, we use normative modelling to investigate sex differences in brain outliers and the influence of *APOE4* status and amyloid centiloid in 372 cognitively normal adults aged 69–75 from the Insight 46 cohort, assessed across two time points. This cohort, consisting of individuals born in 1946, allows for precise control of age effects, and has already shown great value in identifying patterns of amyloid PET changes (Giovane et al., 2025; Lu et al., 2024) and brain-aging trajectories (Subramaniapillai, Schindler, et al., 2024; Wagen et al., 2022). Here, we ran linear mixed models to examine: 1) the effects of age and sex on tOC and PACC scores, 2) the interaction between sex and AD risk (*APOE4* status and amyloid centiloid) on tOC, 3) whether the effects of tOC and amyloid burden on PACC scores vary by sex and *APOE4* status, and 4) the effects of age, sex, and AD risk on longitudinal changes in tOC/PACC.

## Methods

### Participants

The Medical Research Council National Survey of Health and Development (NSHD) is a birth cohort study that has been following 5,362 British individuals since their birth in March 1946. A subgroup from this cohort participated in the neuroscience substudy, Insight 46, where they underwent clinical and cognitive assessments, as well as simultaneous T1-weighted MRI and [^18^F]florbetapir PET imaging using a 3T Siemens Biograph mMR combined PET/MRI scanner. Participants for Insight 46 were randomly selected from those aged 60-64 who had previously agreed to attend a clinic visit in London and had relevant data from childhood and adulthood available. The assessments began in 2015 when the participants were about 70 years old, with timepoint 2 occurring 2 to 4.5 years later, all conducted at a single site. For further details on the study design and recruitment, refer to James et al., 2018; Lane et al., 2017; Mason et al., 2020; Murray-Smith et al., 2024. Table 1 provides an overview of the participant demographics across the two timepoints. See Supplementary Information (SI) Section 1, Figure 1 for a flowchart detailing participant selection and data processing steps.

**Table 1.**
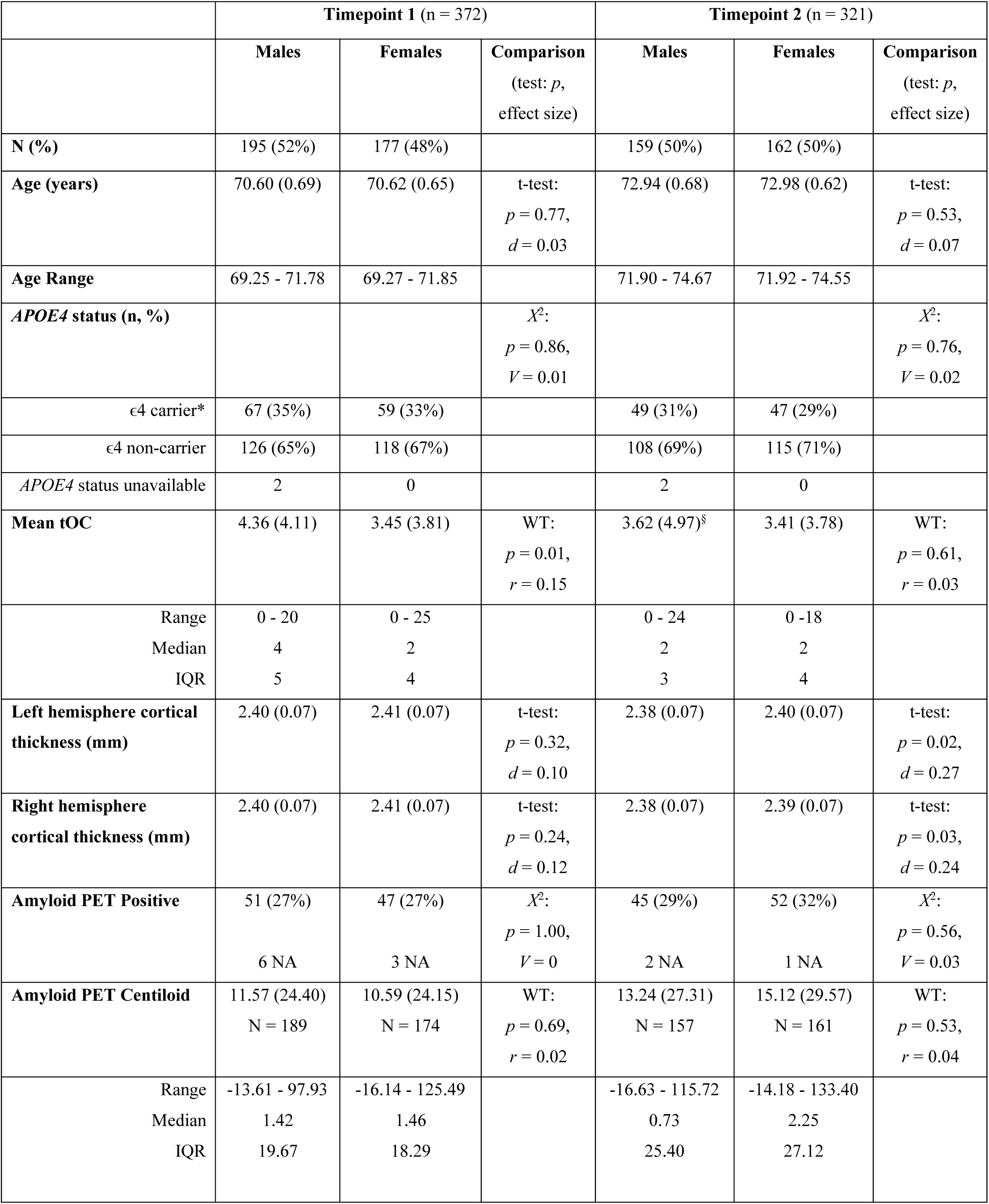
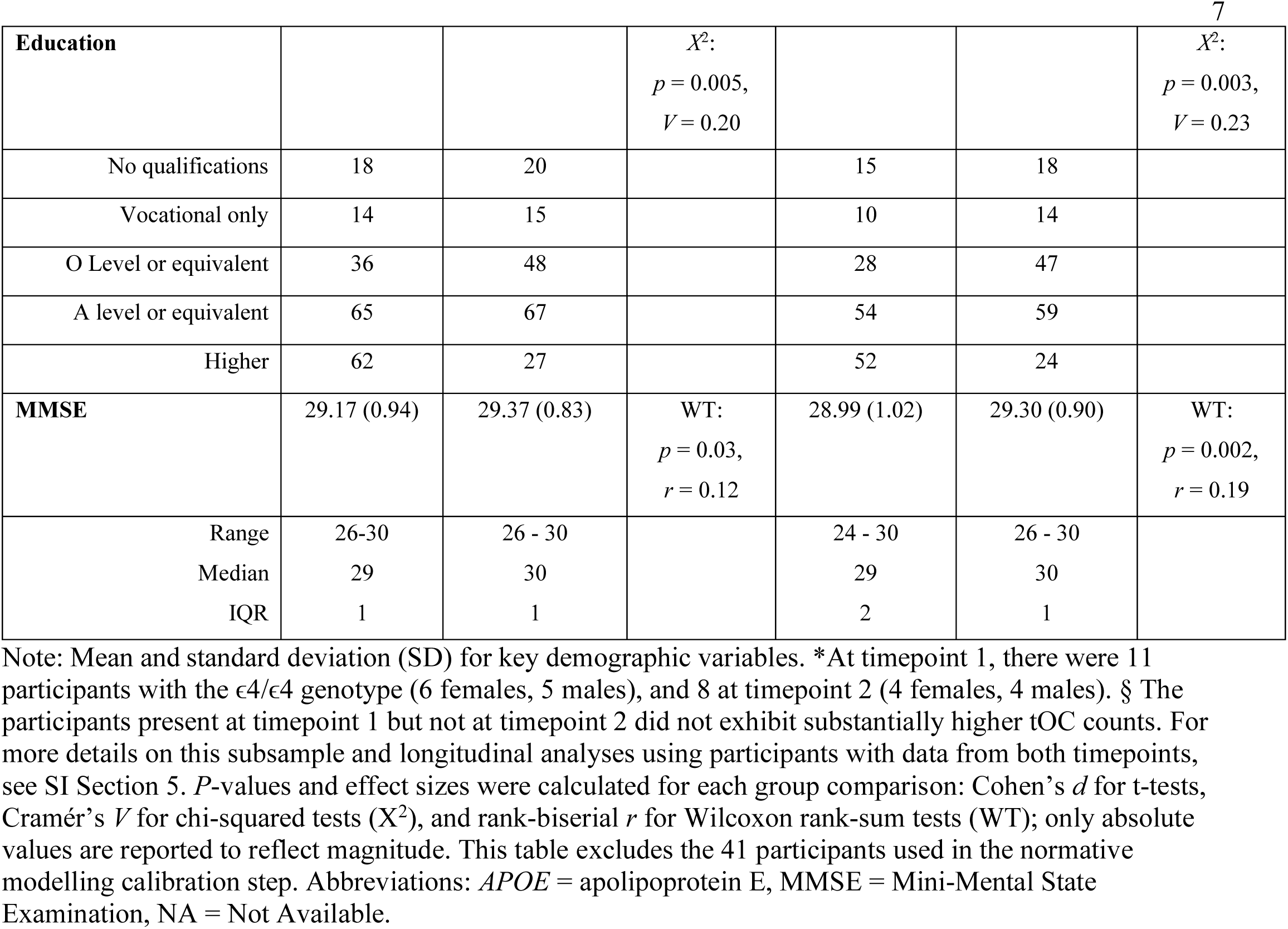
Sample demographics for Insight 46.

### Data Availability

The data supporting the findings of this study were obtained from the MRC National Survey of Health and Development (NSHD). Researchers may request access via the NSHD Data Sharing Committee at https://skylark.ucl.ac.uk/NSHD/doku.php?id=home, subject to institutional approval and data use agreements. The code used for data preprocessing and normative modeling is openly available through the Predictive Clinical Neuroscience Toolkit (PCNtoolkit version 0.20) at https://pcntoolkit.readthedocs.io/en/latest/ and the reference normative models were created using the PCNtoolkit software package (github.com/amarquand/PCNtoolkit).

### APOE genotyping

*APOE*4 carrier status was determined by examining two *APOE* single-nucleotide polymorphisms (rs7412 and rs429358) using TaqMan technology (Lane et al., 2017). Individuals were labelled as “carriers” if they had combinations of ɛ2/ɛ4, ɛ3/ɛ4 and ɛ4/ɛ4, and as “non-carriers” if they had combinations of ɛ2/ɛ2, ɛ2/ɛ3, and ɛ3/ɛ3. Individuals with ɛ4/ɛ4 genotype were not analyzed separately due to the small number of participants across timepoints. The proportion of e4 carriers remained relatively stable at approximately 30% across timepoints 1 and 2 (Table 1).

### Estimation of Amyloid PET Centiloid Values

The standardized uptake value ratios (SUVRs) from [^18^F]florbetapir PET scans were previously processed using a whole cerebellum reference region with partial volume correction (see Coath et al., 2023 for detailed methods). These SUVRs were transformed to the Centiloid (CL) scale to standardize measurements of amyloid PET imaging between radiotracers and pipelines (Coath et al., 2023; Klunk et al., 2015). Amyloid positivity was defined using a bi-modal Gaussian mixture-modelling derived cutpoint of 12 CL, equivalent to the 99th percentile of the lower component.

### Estimation of Cortical Thickness and Subcortical Brain Volumes

T1-weighted MRI data were corrected for gradient non-linearity and manually checked for quality control issues at timepoint 1 and across both time-points. Data that passed quality control were processed cross-sectionally using FreeSurfer version 7.1.0 (Fischl, 2012; Fischl & Dale, 2000), before being processed using the longitudinal stream (Reuter et al., 2012). Cortical thickness segmentations were visually inspected for major errors, and cortical thickness values for 148 regions specified in the Destrieux atlas were extracted (Destrieux et al., 2010). Subcortical brain volume values for 20 regions were also extracted.

### Neuroanatomical Normative Modelling

A hierarchical Bayesian regression model was previously trained on multisite data from 58,836 scans across 82 different sites spanning the human lifespan (ages 2-100) to model regional brain structure (de Boer et al., 2024; Kia et al., 2022; Rutherford et al., 2023; Rutherford, Fraza, et al., 2022; Rutherford, Kia, et al., 2022). The model accounts for age, sex, and site by incorporating them into the predictive framework used to estimate cortical thickness and subcortical volume for each parcel in a high-resolution atlas (Destrieux et al., 2010). We performed a model adaptation step to adjust the model parameters to our specific dataset using a transfer learning approach based on data from 41 cognitively normal Insight 46 participants who only had timepoint 1 data available. This recalibrated model was then applied to the remaining Insight 46 participants, generating regional and mean cortical thickness and subcortical volume Z-scores, which reflect how much an individual’s brain measure differs from what is expected for their age and sex, based on the normative range of the reference dataset.

Z-scores below −1.96, representing the lowest 2.5% of the normal distribution per region, were categorized as outliers in thickness or volume. The focus on these lower thresholds is because they reflect thinning or volume loss, potentially as a result of neurodegeneration (Verdi et al., 2023). The tOC across all 168 regions was calculated by summing the number of brain structure outliers identified for each participant.

### Preclinical Alzheimer Cognitive Composite Scores

The Preclinical Alzheimer Cognitive Composite (PACC) score, already available in the dataset and previously used in several studies (Lane et al., 2019; Mormino et al., 2017; Rabin et al., 2018), was assessed at each timepoint. This score is derived from neuropsychological tests measuring episodic memory (Wechsler Memory Scale-Revised Logical Memory Test [Wechsler, 1987], Face-Name Associative Memory Exam 12 [Papp et al., 2014]), processing speed (Wechsler Adult Intelligence Scale-Revised Digit Symbol Substitution Test [Wechsler, 1981]), and global cognition (Mini-Mental State Examination [Folstein et al., 1975]). Each of the four test components is converted into Z-scores and then averaged, with higher PACC scores indicating better cognitive performance.

### Statistical analyses

All statistical analyses were conducted using R version 4.3. For models with tOC measures as the outcome (i.e., dependent variable), we ran negative binomial generalized linear mixed models (i.e., *glmmTMB* type 2 family) to fit the count data, which exhibited a non-Gaussian distribution (Brooks et al., 2017). We used linear mixed-effects regression models (i.e., *lmer*) for models with PACC scores as the outcome as these continuous cognitive measures approximated a Gaussian distribution (Bates et al., 2015). The precise specification of these models is given below. Sex (2 levels: Male, Female) and *APOE4* status (2 levels: carrier, noncarrier) were treated as categorical variables. Age and centiloid values were standardized (i.e., centered by subtracting the sample mean and scaled by dividing the standard deviation) across the entire dataset and treated as continuous measures. To investigate spatial distributions of brain structure outliers, the percentage of participants within each sex with outliers (i.e., Z scores below −1.96) was determined for each region. This allowed for visualization of the degree of overlap or divergence in regional outlier patterns. Brain surface mapping was performed using the Destrieux (cortical regions) and aseg (subcortical regions) atlas with the R package *ggseg* (Mowinckel & Vidal-Piñeiro, 2019).

#### 1. Associations between age, sex, and tOC/PACC Scores

We first tested the effects of age and sex on tOC and PACC scores:

**Model 1:**

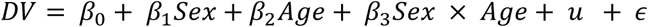

where, DV represents the tOC or PACC scores of participants and *u* represents the modelling of Participant ID as a random intercept, and *ɛ* is the error term. The global intercept is denoted by *β*_0_, while the regression coefficients for *Sex, Age*, and the interaction between *Sex* and *Age* are denoted as *β*_1_, *β*_2_, and *β*_3_ respectively. As sex differences in PACC scores were previously examined in this cohort (Lane et al., 2019; Lu et al., 2019), we uniquely tested whether there were sex differences in the effect of age on PACC scores.

#### 2. Associations between age, sex, AD risk, and tOC/PACC Scores

We then tested whether there were sex differences in the effect of AD risk (*APOE4* genetic risk, centiloid) on tOC across timepoints using the following separate models:

**Model 2:**

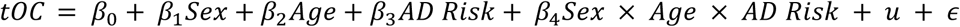

As part of the three-way interaction term, these models test the underlying two-way interactions between sex and AD risk, as well as age and AD risk. As a cross-check, we also ran separate models testing for two-way interactions only (provided in the supplementary material). To determine whether the effect of greater brain pathology, from brain outliers and amyloid accumulation, on cognitive scores varied by sex, we ran the following model to examine the interaction between sex, centiloid, and tOC on PACC scores, with participants’ age at the time of the scan included as a covariate:

**Model 3:**

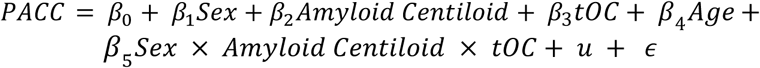

For model 3, we ran stratified analyses by *APOE4* status to assess whether sex differences in the relationship between brain pathology and cognition varied by genetic risk for AD.

#### 3. Associations between age, sex, and AD risk on longitudinal tOC/PACC changes

We performed simple linear regressions to explore the associations between age, sex, AD risk and longitudinal changes in tOC/PACC in a subsample of participants with data from both time points (n = 309; see SI Section 1, Table 1 for demographics of this sample). We calculated change using difference scores [Timepoint 2 values − Timepoint 1 values], and included assessment interval as a covariate so that observed changes in tOC are not biased by differences in time between assessments (mean assessment interval = 2.4 years, SD = 0.2, range = [2.0 to 3.51]). Difference scores offer a straightforward measure of individual change over time (Gollwitzer et al., 2014; McArdle, 2009), and have previously been used in normative modelling of a longitudinal cohort (Verdi et al., 2024).

**Model 4:**

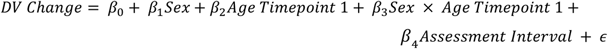

**Model 5:**

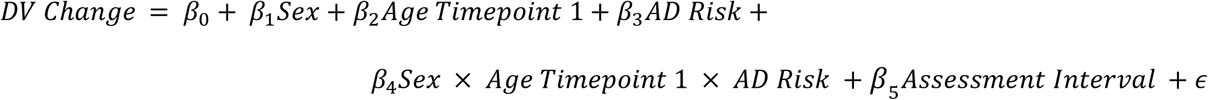

In model 5, we used amyloid PET status (negative or positive) at each timepoint to create a new variable that categorizes participants by changes in amyloid positivity over time. This variable has three possible values: 0 for participants who were amyloid negative at both timepoints (n = 212), 1 for those with amyloid present at both timepoints (n = 70), and 2 for participants who had converted to amyloid positivity status from timepoint 1 to timepoint 2 (n = 19; converters). There were no reverters from positive to negative in this dataset. To test if the results differed when using an alternative measure of change in tOC/PACC, we reran supplementary analyses of models 4 and 5 using timepoint 2 values with timepoint 1 values regressed out (i.e., modelling changes in DV independent of its initial value; Gollwitzer et al., 2014). This approach can be useful when timepoint 1 tOC/PACC values vary substantially between participants, as the residualized change helps account for timepoint 1 differences (Vickers & Altman, 2001).

We conducted supplementary analyses across all models using a matched longitudinal sample of participants with data available at both timepoints to assess whether the observed effects persist within the same individuals over time.

## Results

### 1. Associations between age, sex, and tOC/PACC Scores

Model 1 revealed a significant interaction between age and sex on tOC (β = −0.13, SE = 0.05, z = −2.39, p = 0.02), indicating that males had higher tOC than females at younger ages. PACC scores were greater in females compared to males (β = −0.33, SE = 0.06, t = −5.26, p < 0.005). There was no significant effect of age (β = −0.02, SE = 0.02, t = −1.40, p = 0.16) or interaction between age and sex on PACC (β = 0.02, SE = 0.02, t = 0.84, p = 0.40). SI Section 2, Figure 2 shows the relationships between age and tOC and PACC scores in females and males.

Figures 1–2 show the proportion of participants with outliers in each brain region, for females and males at each timepoint. While tOC captures the total number of outlier regions per individual, these maps reflect group-level variation across regions, highlighting how the spatial distribution of outliers differs within and between groups. For males at timepoint 1, regions most affected were left middle occipital and lunatus sulci (35.38%), right superior transverse occipital sulcus (26.15%), right middle occipital and lunatus sulci (21.54%), left superior transverse occipital sulcus (19.49%), right long insular gyrus and central sulcus of the insula (10.26%), and right superior part of the precentral sulcus (9.23%). For females at timepoint 1, the regions with the highest outlier percentage were left superior transverse occipital sulcus (14.69%), left suborbital sulcus (9.04%), left posterior-dorsal part of the cingulate gyrus (8.00%), left anterior vertical ramus of the lateral fissure (8.00%), left intraparietal and transverse parietal sulci (8.00%), and left medial orbital sulcus and olfactory sulcus (8.00%).

**Figure 1.**
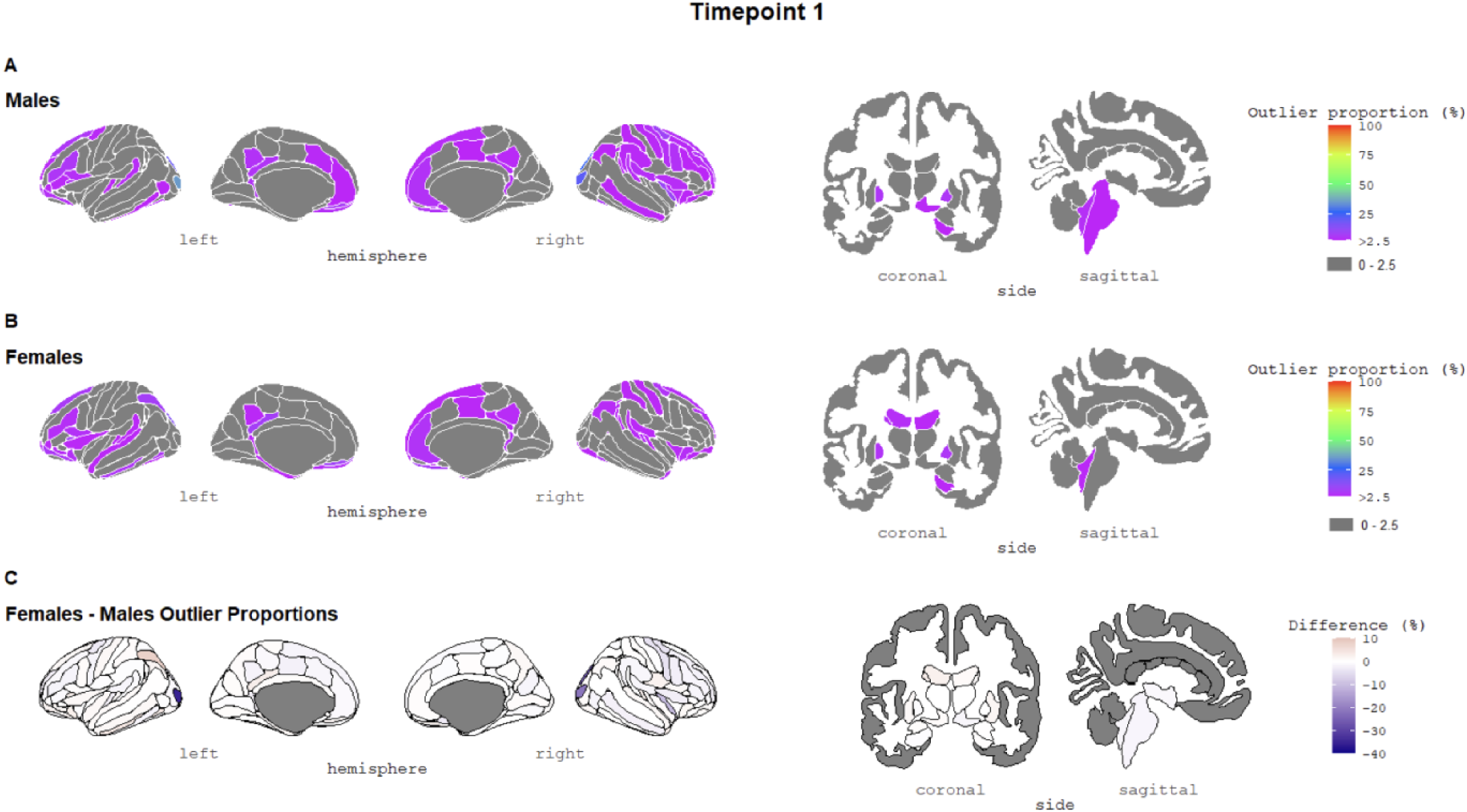
The percentage of outliers present within (A) males and (B) females at timepoint 1. The colour bar represents the outlier proportion (thresholding of z-scores). Regions in grey reflect areas where participants did not have any outliers (≤ 2.5 %). (C) The difference in outlier proportions between females and males. The colour gradient indicates the direction and magnitude of the difference: dark blue represents regions where males have more outliers than females, white indicates no difference between sexes, and dark red represents regions where females have more outliers than males. Colour scales are adjusted to reflect the range of group differences at each timepoint. See SI Figure 3 for an outlier map with the colour scale truncated to enhance visibility of regional variations.

**Figure 2.**
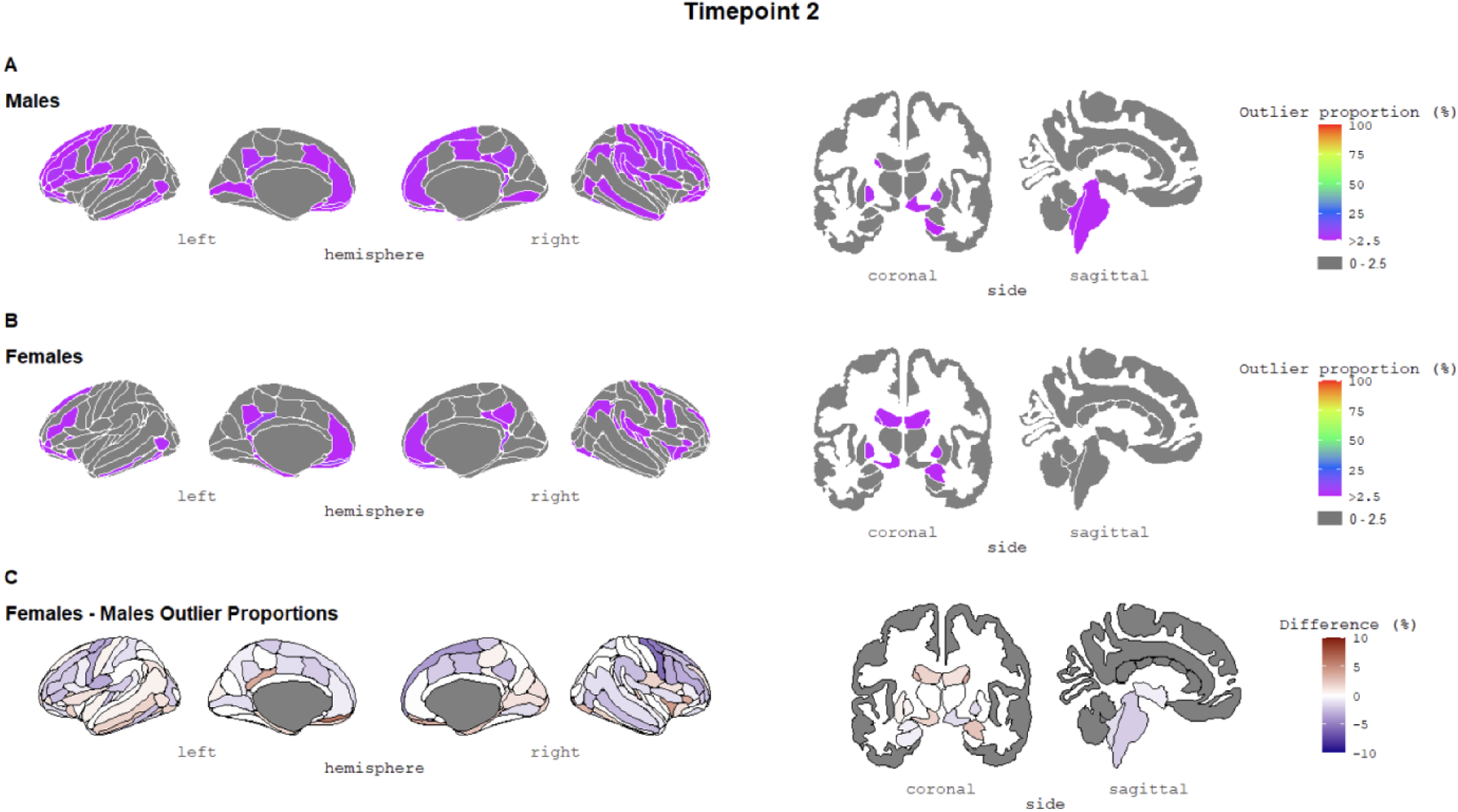
The percentage of outliers present within (A) males and (B) females at timepoint 2. The colour bar represents the outlier proportion (thresholding of z-scores). Regions in grey reflect areas where participants did not have any outliers (≤ 2.5 %). (C) The difference in outlier proportions between females and males. The colour gradient indicates the direction and magnitude of the difference: dark blue represents regions where males have more outliers than females, white indicates no difference between sexes, and dark red represents regions where females have more outliers than males. Colour scales are adjusted to reflect the range of group differences at each timepoint. At timepoint 1, group differences were larger but observed in fewer regions; at timepoint 2, differences were smaller in magnitude but present across more regions, resulting in higher visual contrast under the narrower scale. See SI Figure 4 for an outlier map with the colour scale truncated to enhance visibility of regional variations.

For males at timepoint 2, regions most affected were right superior part of the precentral sulcus (10.06%), left medial orbital sulcus and olfactory sulcus (8.81%), right inferior part of the precentral sulcus (8.18%), and right lateral occipitotemporal sulcus (7.55%). For females at timepoint 2, the regions with the highest outlier percentage were left suborbital sulcus (10.49%), left posterior-dorsal part of the cingulate gyrus (9.26%), right gyrus rectus (8.02%), right posterior-ventral part of the cingulate gyrus (7.40%), and left medial orbital sulcus and olfactory sulcus (7.40%).

### 2. Associations between age, sex, AD risk, and tOC

Model 2 showed no significant three-way interaction between age, sex, and *APOE4* status on tOC (β = 0.13, SE = 0.12, z = 1.14, p = 0.26), nor between age, sex, and centiloid on tOC (β = −0.02, SE = 0.05, z = −0.36, p = 0.72). See SI Section 3, Tables 2-7 for underlying two-way interactions and separate regressions with two-way interactions.

#### 2.1 Associations between sex, amyloid burden, and tOC on cognitive outcomes

Model 3 revealed a significant three-way interaction between sex, centiloid, and tOC on PACC scores (β = 0.11, SE = 0.04, t = 2.52, p = <0.05; SI Section 3, Figure 5). In females, greater amyloid burden was associated with poorer cognitive performance at higher tOC levels (β = −0.10, SE = 0.04, p = 0.007), whereas this association was not observed in males (β = 0.01, SE = 0.02, p = 0.69; based on the model re-estimation with male as the reference category). When this model was run within *APOE4* non-carriers, no significant three-way interaction was observed (β = 0.04, SE = 0.09, t = 0.41, p = 0.68). However, among *APOE4* carriers, females with both greater tOC and amyloid burden showed worse cognitive outcomes (β = −0.11, SE = 0.05, p = 0.04), whereas this association was not observed in male *APOE4* carriers (β = 0.005, SE = 0.03, p = 0.86). Figure 3 illustrates the relationship between tOC and PACC scores in *APOE4* non-carriers and carriers. See SI Section 3, Tables 8-19 for two-way interactions and stratified regression results.

**Figure 3.**
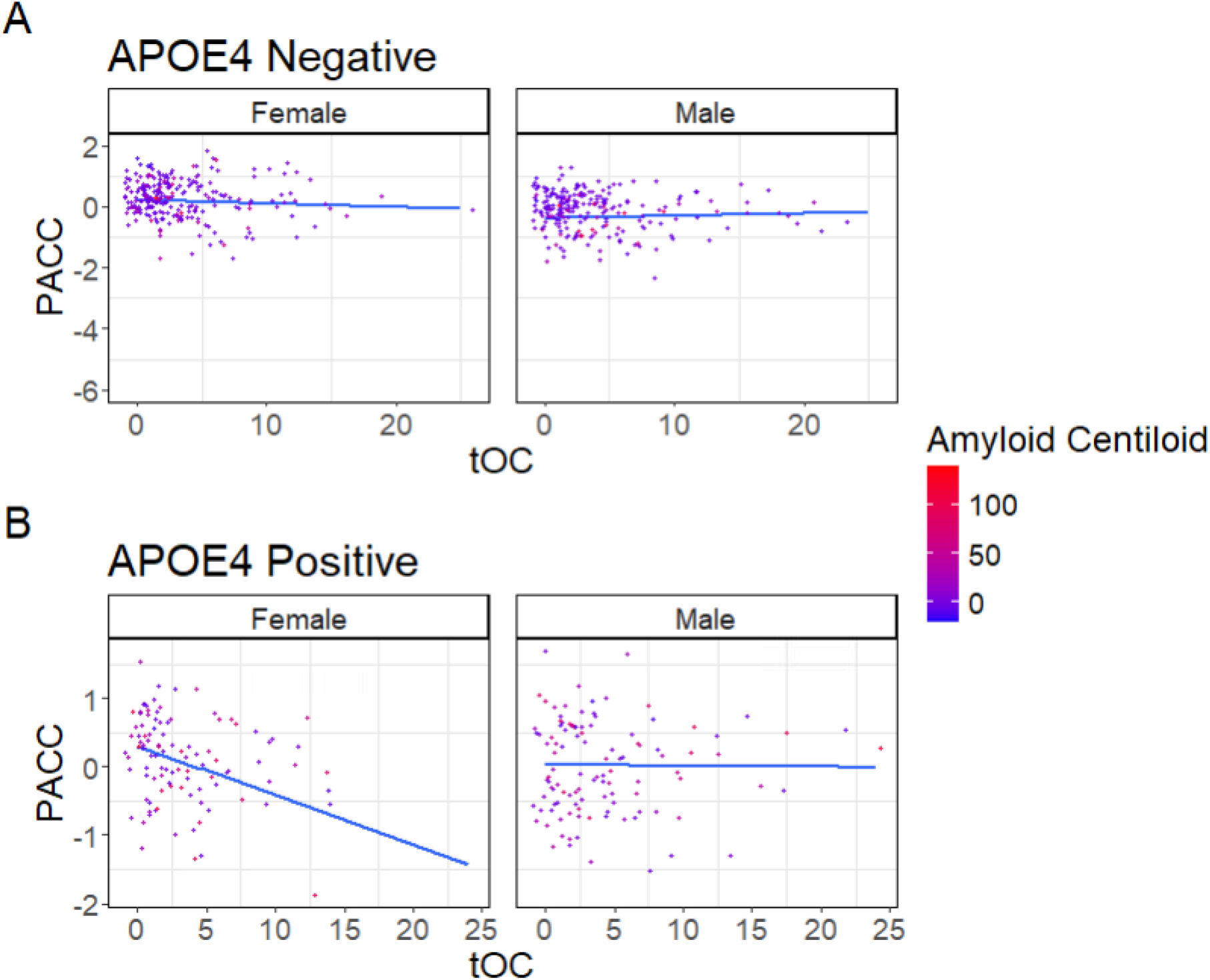
Fitted linear relationships between tOC and PACC scores in *APOE4* (A) non-carriers and (B) carriers, with their corresponding 95% confidence intervals (model 3). Data points on the plots represent raw data from the dataset and are colour-coded based on amyloid centiloid. Abbreviations: PACC = Preclinical Alzheimer Cognitive Composite; tOC = total outlier count; *APOE4* = apolipoprotein E4 status.

### 3. Associations between age, sex, and AD risk on longitudinal tOC/PACC changes

Model 4 revealed a significant interaction between age and sex on changes in tOC (β = 1.89, SE = 0.34, t = 5.58, p = <0.01), indicating that an older age was related to a greater increase in tOC, with stronger effects in males compared to females (Figure 4). When changes in PACC scores were used as the outcome, this model did not reveal any significant effects of age (β = −0.01, SE = 0.03, t = −0.41, p = 0.69), sex (β = 0.01, SE = 0.05, t = 0.33, p = 0.74), or their interaction (β = 0.04, SE = 0.05, t = 0.86, p = 0.39; Figure 4).

**Figure 4.**
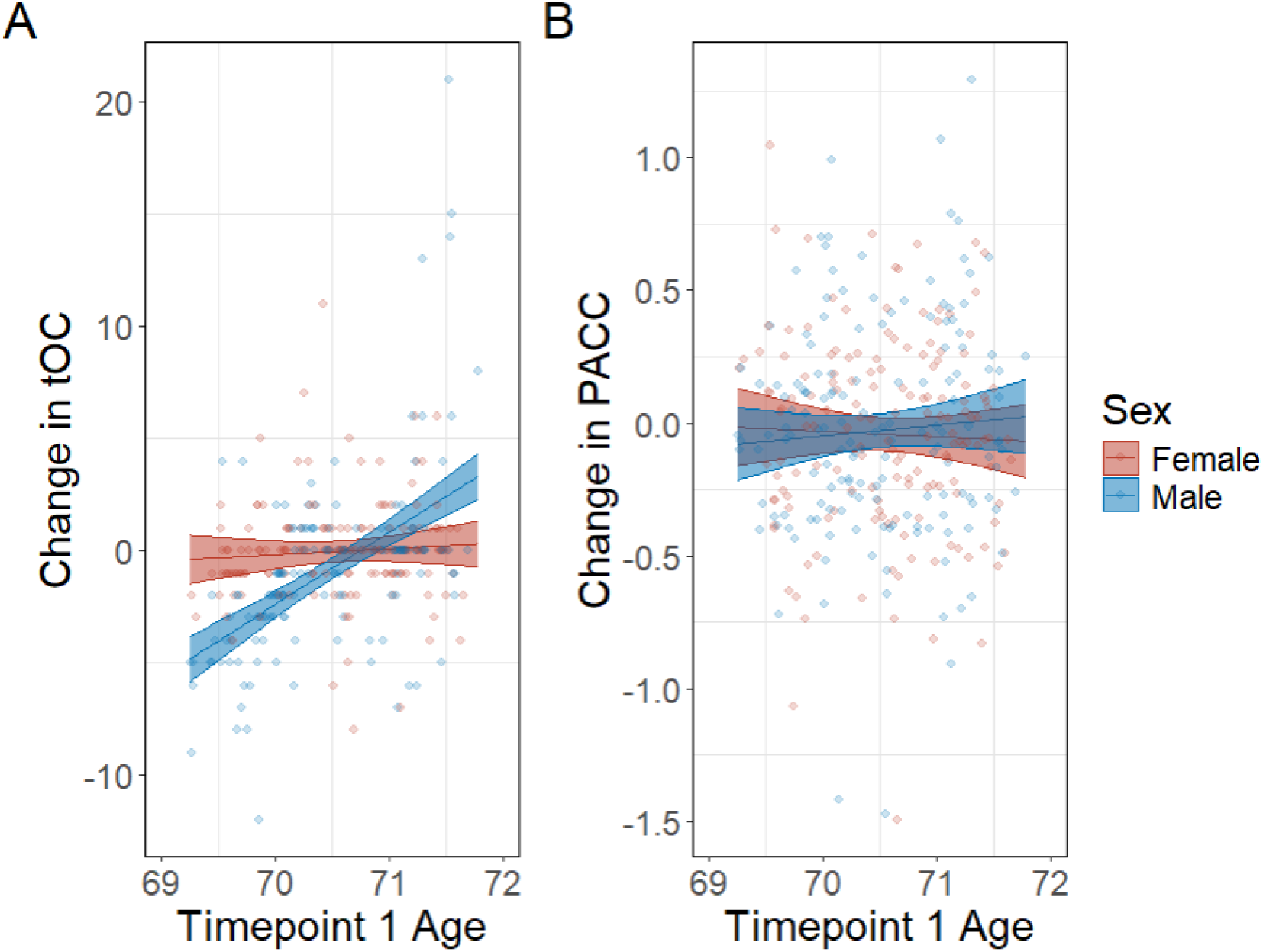
Fitted relationships between age at timepoint 1 and changes in tOC (A) or PACC (B), with corresponding 95% confidence intervals (model 4). Change was measured using difference scores [Timepoint 2 values − Timepoint 1 values]. Data points on the plots represent raw data from the dataset, while lines of best fit and confidence intervals are model derived. Abbreviations: PACC = Preclinical Alzheimer Cognitive Composite; tOC = total outlier count.

Model 5 showed no significant interactions between age, sex, and *APOE4/*amyloid status on changes in tOC/PACC. Refer to SI Section 3, Tables 20-23 for a detailed list of regression outputs from these models. The supplementary change score analyses of models 4 and 5, which regressed out timepoint 1 values to account for baseline differences in tOC/PACC, revealed findings consistent with the primary analyses (see SI Section 4). The supplementary analyses using the matched longitudinal sample were consistent across models with those from the primary analyses (see SI Section 5).

## Discussion

This longitudinal birth-cohort study highlights variation in brain and cognitive patterns across sex and AD risk groups among cognitively normal older adults. In summary, males showed more brain structural outliers per individual than females, primarily at younger ages. At timepoint 1, males had outliers in more regions, although high outlier proportions were mostly confined to occipital areas. At timepoint 2, group differences became more spatially distinct, as males and females showed outliers in different brain regions. Older males exhibited steeper increases in brain structural outliers over time, as reflected in an age-by-sex interaction on longitudinal tOC change. Females showed higher PACC scores overall, but we found no sex differences in cognitive change over time. Greater tOC and amyloid burden were both associated with poorer cognitive scores cross-sectionally, with the strongest associations observed in female *APOE4* carriers. However, we found no evidence that AD risk influenced age-related changes in tOC or cognition over the follow-up period. Together, these findings point to subtle but distinct brain and cognitive patterns across sex and AD risk groups, offering insight into early variation that may precede disease-related decline.

In our sample, cross-sectional sex differences in tOC were evident primarily at younger ages, suggesting that males may experience earlier structural brain vulnerability. For example, previous studies have shown that males exhibit steeper age-related declines in cortical thickness and gray matter volume beginning in midlife (Driscoll et al., 2009; Pacheco et al., 2015; Thambisetty et al., 2010), potentially driven by faster vascular changes (Nowell et al., 2025). However, our findings may also reflect selective attrition, as more males dropped out by timepoint 2. Prior research on this cohort has shown that dropout was more likely among individuals with lower cognitive performance and socioeconomic status (James et al., 2018). While this could result in a healthier remaining male subsample that attenuated group-level differences, supplementary analyses using only participants with data from both timepoints showed consistent results. In addition, participants who were present only at timepoint 1 did not exhibit substantially higher tOC, suggesting that attrition was not driving the primary findings. Our findings align with previous research reporting older brain age estimates and poorer gray matter health in males compared to females in cognitively normal older adult cohorts, including studies using this sample (Armstrong et al., 2019; Cole et al., 2018; Cowell et al., 1994; Driscoll et al., 2009; Subramaniapillai, Schindler, et al., 2024; Wagen et al., 2022). Our longitudinal analyses further revealed that males who were older at timepoint 1 showed greater increases in total brain outlier count over time compared to females, indicating steeper structural decline with advancing age. While the short follow-up window limits interpretation of long-term trajectories, this suggests that sex differences in the timing of structural brain decline may emerge even within a narrow age range in later life.

Although all participants in this sample were cognitively normal, the spatial patterns of brain deviation overlap with regions commonly implicated in the early stages of AD. Regions more affected in males, such as the occipital sulci, cingulate cortex, and precentral sulcus, are associated with visuospatial processing, attention, and sensorimotor integration, and have been linked to atypical or early-onset AD presentations (Javitt et al., 2023; Quental et al., 2009). In females, higher outlier proportions at were observed in regions such as the cingulate cortex, gyrus rectus, and olfactory areas, which are also frequently implicated in early AD, particularly in relation to memory and default mode network function (Leech & Sharp, 2014; Mevel et al., 2011). Taken together, these findings suggest that even among cognitively normal individuals, sex differences in the spatial distribution of brain deviation may reflect distinct pathways of brain aging (Sanford et al., 2022). While the regions involved differ between sexes, both sets include areas that are vulnerable in preclinical AD, pointing to sex-specific trajectories that may have relevance for early detection efforts. Given the overlap between regions involved in normal aging and those affected in early AD (Fjell et al., 2014; Roe et al., 2024), future work is needed to distinguish patterns that reflect early signs of AD processes. Importantly, group-average outlier maps reveal regional trends but can obscure substantial inter-individual variability. Here, the highest proportion of outliers were observed in males, but only four regions showed substantially higher values of overlap between participants (20-35%), while the overlap in the remaining regions was comparable to females. This suggests that the higher male tOC at timepoint 1 was driven by a small subset of regions with more consistent deviations, rather than widespread differences across the brain. If atrophy were homogenous, we would expect near-complete overlap (i.e., 100% of individuals with outliers in the same region). Instead, the spatial distribution of deviations was heterogeneous, underscoring the need for individual-level analyses. This is consistent with previous studies in AD-diagnosed samples, which have similarly highlighted considerable variability in spatial atrophy patterns (Loreto et al., 2024; Verdi et al., 2023, 2024).

Our findings align with this sample’s previously established female advantage on PACC scores (Lane et al., 2019; Lu et al., 2019). Importantly, the PACC score has a strong verbal memory component, with females typically outperforming males, which may make it more challenging to detect early signs of neurodegeneration in females (Emrani & Sundermann, 2025). When we stratified the data by both sex and AD risk, we detected lower cognitive performance in female *APOE4* carriers with higher levels of amyloid and outlier counts compared to males with the same level of risk. Although we did not observe a statistical interaction between sex and age on cognitive change, suggesting that cognitive performance did not significantly differ over time between sexes, this may change when pathological burden thresholds are met. Prior research suggests that sex differences in pathology and cognition tend to be more pronounced at later disease stages (Barnes et al., 2005; Vila-Castelar et al., 2025; Zhu et al., 2021). However, recent studies have demonstrated cognitive decline over a three-year timeframe in cognitively normal individuals with elevated amyloid loads, with an age range similar to that of our current sample (Quenon et al., 2024). While cross-sectional findings may partly reflect pre-existing group differences (Karama et al., 2014; Richards et al., 2019), our results might capture early cognitive vulnerability in females with elevated AD risk that would have gone unnoticed without stratified analyses. A recent study from the same cohort showed that greater Aβ deposition and brain pathology predicted faster rates of preceding cognitive decline (James et al., 2023). This underscores the importance of tracking participants over longer periods of time, to better understand when and how sex differences in brain pathology and cognitive decline emerge.

Our white, UK-based sample with high educational attainment may have limited generalizability to more diverse and socioeconomically varied groups (Keuss et al., 2022). Although this aligns with the normative modelling approach, which is typically trained on relatively homogenous datasets and thus may have reliable model generalizability, it still limits the applicability of our findings to more diverse populations. Additionally, sex-specific factors, such as menopause and hormone therapy use, may offer valuable insights into the differences between males and females in brain aging (Needham et al., 2023). These factors may interact with cardiometabolic risk and inflammation (Caldwell et al., 2021; Fatih et al., 2022), influencing brain health in ways that differ by sex (Nowell et al., 2025; Subramaniapillai et al., 2022). Future research should prioritize the inclusion of underrepresented groups and broader risk profiles to better capture the complexities of sex- and gender-related factors in brain aging and disease risk (Arenaza-Urquijo et al., 2024; Bonkhoff et al., 2025; Subramaniapillai et al., 2024).

In summary, our findings reveal a nuanced picture of brain and cognitive health in older adulthood, shaped by individual differences such as sex and AD risk. Although males showed greater brain outliers than females at younger ages, outlier maps highlighted greater sex-specific heterogeneity in the spatial distribution of structural deviations at follow-up. Importantly, stratified analyses revealed that the overall female advantage in cognitive performance may mask early cognitive deficits in at-risk females. The observed effects, though small, underscore the need for further research with more diverse and representative longitudinal samples to better characterize sex differences in brain health and AD risk with age.

## Acknowledgements

The authors received funding from the Swiss National Science Foundation (PZ00P3_193658 to AMdL; TMPFP3_217174 to SS); the Natural Sciences and Engineering Research Council of Canada (Postdoctoral Fellowship Grant to SS). FB is supported by the NIHR biomedical research centre at UCLH. JHC and SV are supported by the Alzheimer’s Society (grant reference 634). JMS is a National Institute for Health Research (NIHR) Senior Investigator and acknowledges the support of the NIHR University College London Hospitals Biomedical Research Centre and the UCL Centre of Research Excellence, an initiative funded by British Heart Foundation (RE/24/130013). He has grant funding from Alzheimer’s Research UK, Brain Research UK, Weston Brain Institute, Medical Research Council, British Heart Foundation, Wolfson Foundation, UK Dementia Research Institute and Alzheimer’s Association. This work uses data provided by study participants or patients and collected through clinical studies or as part of their care and support.

Insight 46 is funded by grants from Alzheimer’s Research UK (ARUK-PG2014-1946, ARUK-PG2017-1946 principal investigators (PIs) JMS, NCF, MR), Alzheimer’s Association (SG-666374-UK BIRTH COHORT PI JMS), the Medical Research Council Dementias Platform UK (CSUB19166 PIs JMS, NCF, MR), The Wolfson Foundation (PR/ylr/18575 PIs NCF, JMS), The Medical Research Council (MC-UU-12019/1 PI MR, MC-UU-12019/3 PI MR), Selfridges Group Foundation (22/3/18 PIs JMS) and Brain Research Trust (UCC14191, PI JMS).

