## Supplement for "Mapping sex differences in brain and cognition in relation to *APOE4* and amyloid burden: A longitudinal normative modelling study"

**Supplementary Information (SI)**

**Section 1. Demographics of Insight 46 Subsample with Two Timepoints**

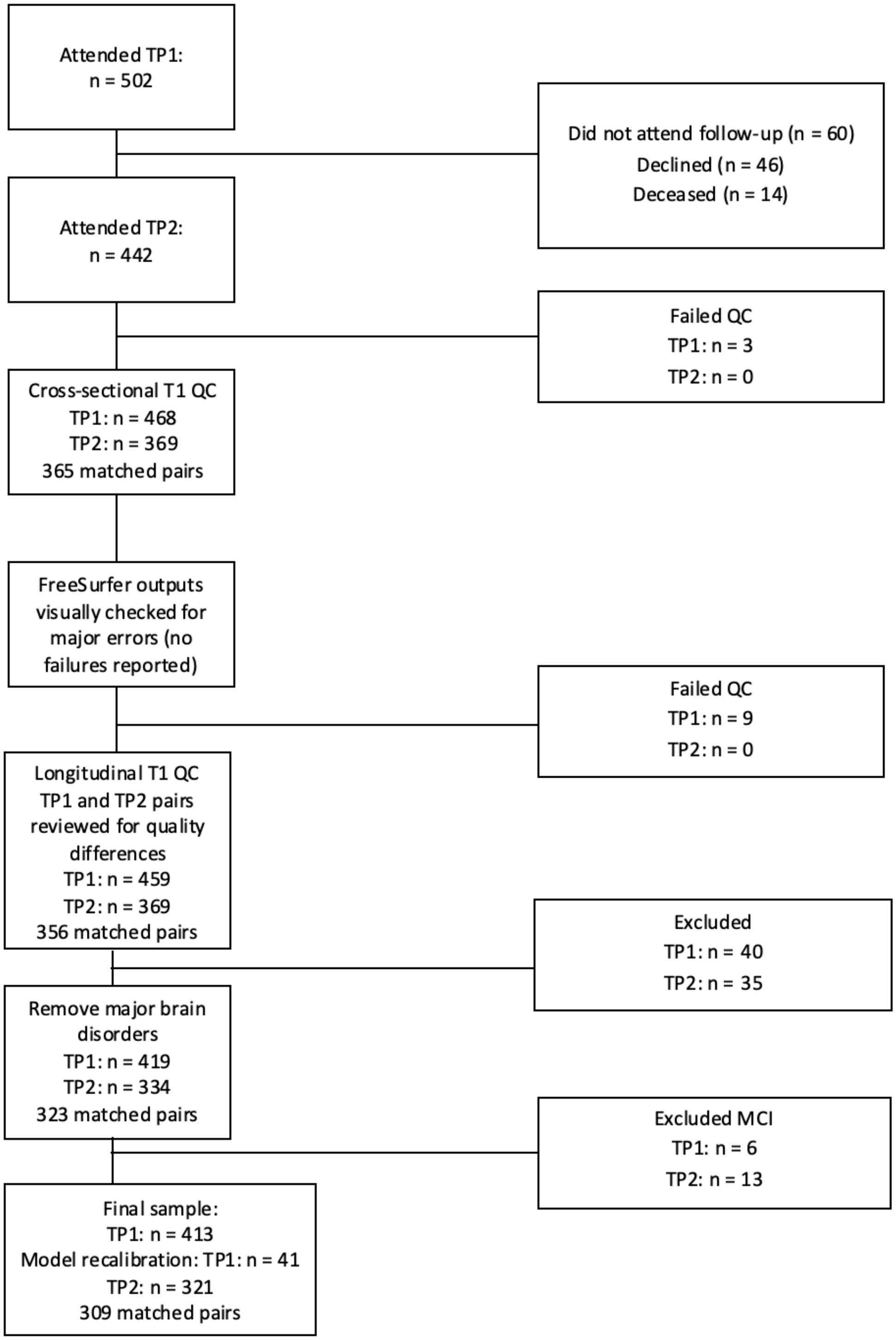


**SI Figure 1.** Flowchart detailing participant selection and data processing steps. Note: TP1 = Timepoint 1; TP2 = Timepoint 2

**SI Table 1.** Sample demographic for Insight 46 sample with information at two timepoints

| | **Timepoint 1** (n = 309) | | | **Timepoint 2** (n = 309) | | |
|---|---|---|---|---|---|---|
| | **Males** | **Females** | **Comparison** (test: *p*, effect size) | **Males** | **Females** | **Comparison** (test: *p*, effect size) |
| **N (%)** | 154 (50%) | 155 (50%) | | 154 (50%) | 155 (50%) | |
| **Age (years)** | 70.50 (0.65) | 70.56 (0.63) | t-test: $p = 0.88$, $d = 0.10$ | 72.93 (0.68) | 72.98 (0.62) | t-test: $p = 0.51$, $d = 0.07$ |
| **Age range** | 69.25 - 71.78 | 69.27 - 71.66 | | 71.90 - 74.67 | 71.92 - 74.55 | |
| ***APOE4* status (n, %)** | | | $X^2$: $p = 0.81$, $V = 0.01$ | | | $X^2$: $p = 0.81$, $V = 0.01$ |
| e4 carrier | 47 (31%) | 45 (29%) | | 47 (31%) | 45 (29%) | |
| e4 non-carrier | 105 (69%) | 110 (71%) | | 105 (69%) | 110 (71%) | |
| *APOE4* status unavailable | 2 | 0 | | 2 | 0 | |
| **Mean tOC** | 4.41 (4.09) | 3.39 (3.83) | WT: $p = 0.01$, $r = 0.17$ | 3.61 (4.95) | 3.38 (3.84) | WT: $p = 0.80$, $r = 0.02$ |
| Range<br>Median<br>IQR | 0-20<br>4<br>5 | 0-25<br>2<br>4 | | 0-24<br>2<br>3 | 0-18<br>2<br>4 | |
| **Left hemisphere cortical thickness (mm)** | 2.40 (0.07) | 2.41 (0.07) | t-test: $p = 0.19$, $d = 0.15$ | 2.38 (0.07) | 2.40 (0.07) | t-test: $p = 0.03$, $d = 0.25$ |
| **Right hemisphere cortical thickness (mm)** | 2.40 (0.07) | 2.41 (0.07) | t-test: $p = 0.16$, $d = 0.16$ | 2.38 (0.07) | 2.39 (0.07) | t-test: $p = 0.05$, $d = 0.23$ |
| **Amyloid PET Positive** | 36 (24%)<br>2 NA | 35 (23%)<br>3 NA | $X^2$: $p = 1.00$, $V = 0$ | 44 (29%)<br>2 NA | 49 (32%)<br>1 NA | $X^2$: $p = 0.18$, $V = 0.02$ |
| **Amyloid PET Centiloid** | 9.91 (23.07)<br>N = 152 | 8.55 (22.04)<br>N = 152 | WT: $p = 0.62$, $r = 0.17$ | 13.52 (27.39)<br>N = 152 | 14.73 (29.04)<br>N = 154 | WT: $p = 0.65$, $r = 0.02$ |
| Range<br>Median<br>IQR | -13.61 - 97.93<br>1.07<br>13.79 | -16.14 - 96.94<br>0.91<br>13.42 | | -16.63 - 115.72<br>1.07<br>25.23 | -14.18 - 133.40<br>2.56<br>26.29 | |

| | | | | | | |
|---|---|---|---|---|---|---|
| **Education** | | | $X^2$: $p = 0.01$, $V = 0.21$ | | | $X^2$: $p = 0.01$, $V = 0.21$ |
| No qualifications | 14 | 17 | | 14 | 17 | |
| Vocational only | 10 | 14 | | 10 | 14 | |
| O Level or equivalent | 27 | 44 | | 27 | 44 | |
| A level or equivalent | 53 | 56 | | 53 | 56 | |
| Higher | 50 | 24 | | 50 | 24 | |
| **MMSE** | 29.23 (0.87) | 29.43 (0.76) | WT: $p = 0.04$, $r = 0.12$ | 28.99 (1.04) | 29.32 (0.90) | WT: $p = 0.002$, $r = 0.19$ |
| Range<br>Median<br>IQR | 26 - 30<br>29<br>1 | 27 - 30<br>30<br>1 | | 24 - 30<br>29<br>2 | 26 - 30<br>30<br>1 | |

Note: Mean and standard deviation (SD) for key demographic variables. *P*-values and effect sizes were calculated for each group comparison: Cohen's *d* for t-tests, Cramér's *V* for chi-squared tests ($X^2$), and rank-biserial *r* for Wilcoxon rank-sum tests (WT); only absolute values are reported to reflect magnitude. This table excludes the 41 participants used in the normative modelling calibration step. Abbreviations: *APOE* = apolipoprotein E, MMSE = Mini-Mental State Examination, NA = Not Available.

**Section 2. Relationships Between Age, Sex, and tOC/PACC Across Timepoints**

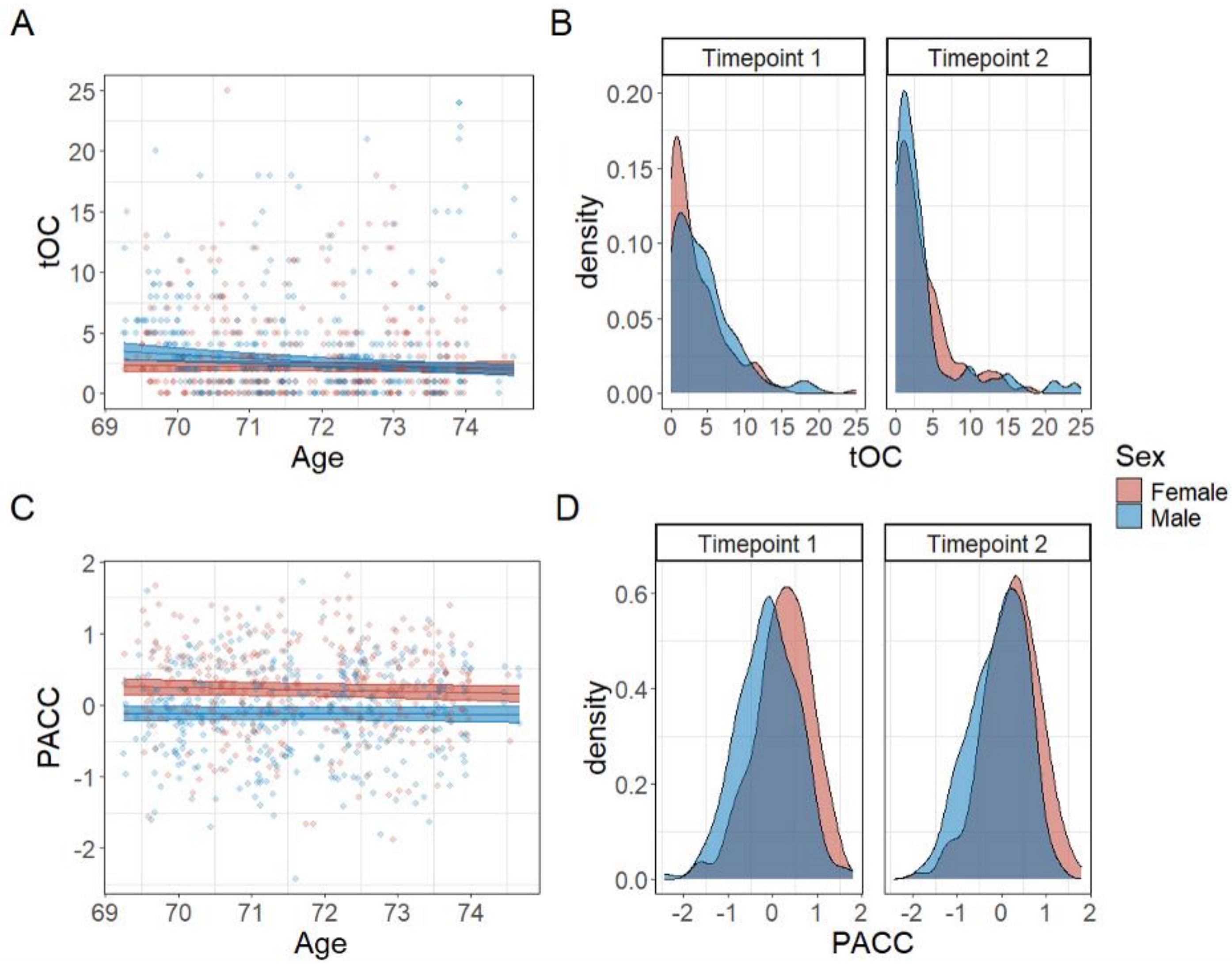


**SI Figure 2**. Fitted relationships between age across both timepoints and tOC (A) and PACC scores (C) in females (red) and males (blue), with their corresponding 95% confidence intervals. Data points on the plots represent raw data from the dataset. Density plots illustrating the distribution of tOC (B) and PACC (D) values, stratified by sex and timepoint. Abbreviations: tOC = total outlier count; PACC = Preclinical Alzheimer Cognitive Composite.

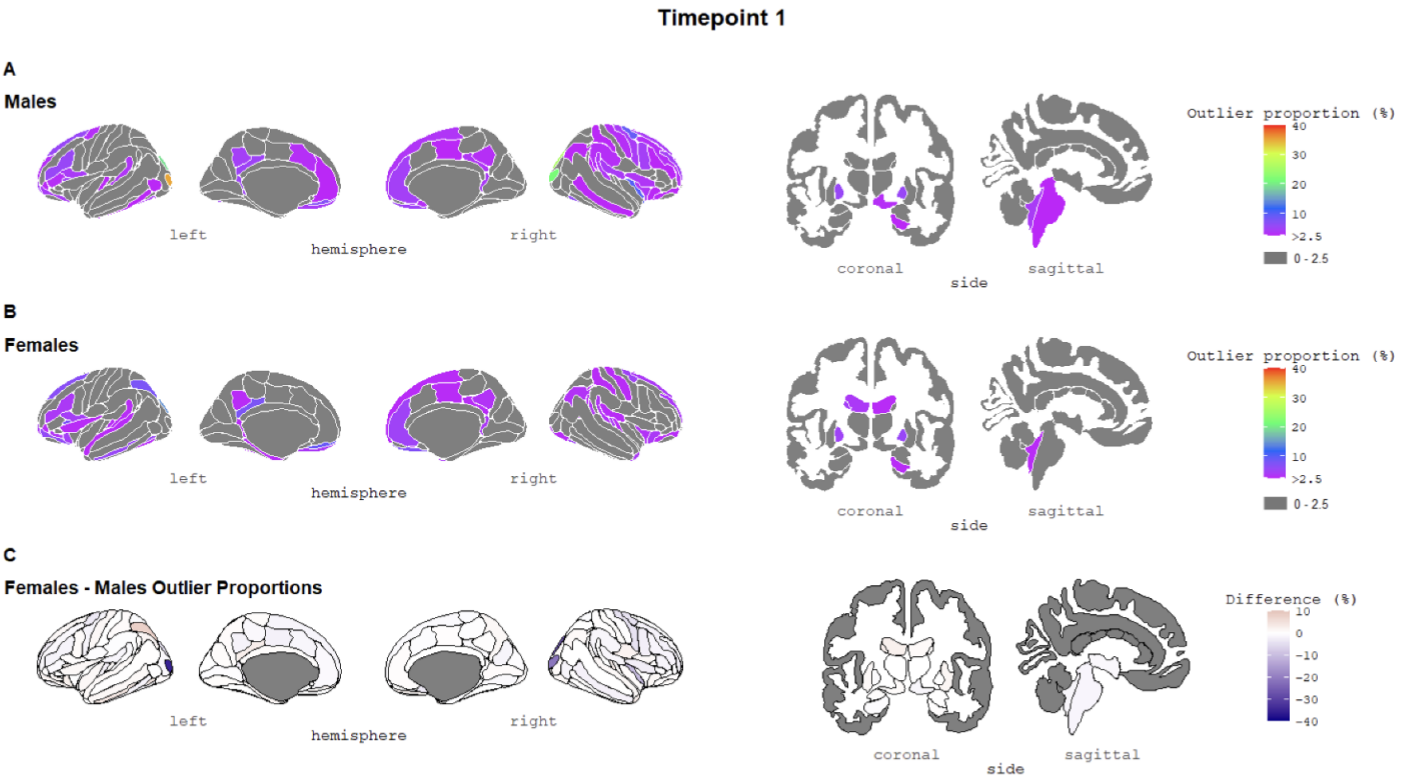


**SI Figure 3.** The percentage of outliers present within (A) females and (B) males at timepoint 1. The colour bar represents the outlier proportion (thresholding of z-scores). Regions in grey reflect areas where participants did not have any outliers (≤ 2.5 %). (C) The difference in outlier proportions between females and males. The colour gradient indicates the direction and magnitude of the difference: dark blue represents regions where males have more outliers than females, white indicates no difference between sexes, and dark red represents regions where females have more outliers than males. Color scale truncated at 40% to enhance visibility of regional variation. Same data visualized with full 0–100% scale for reference in Figure 1 of the main manuscript.

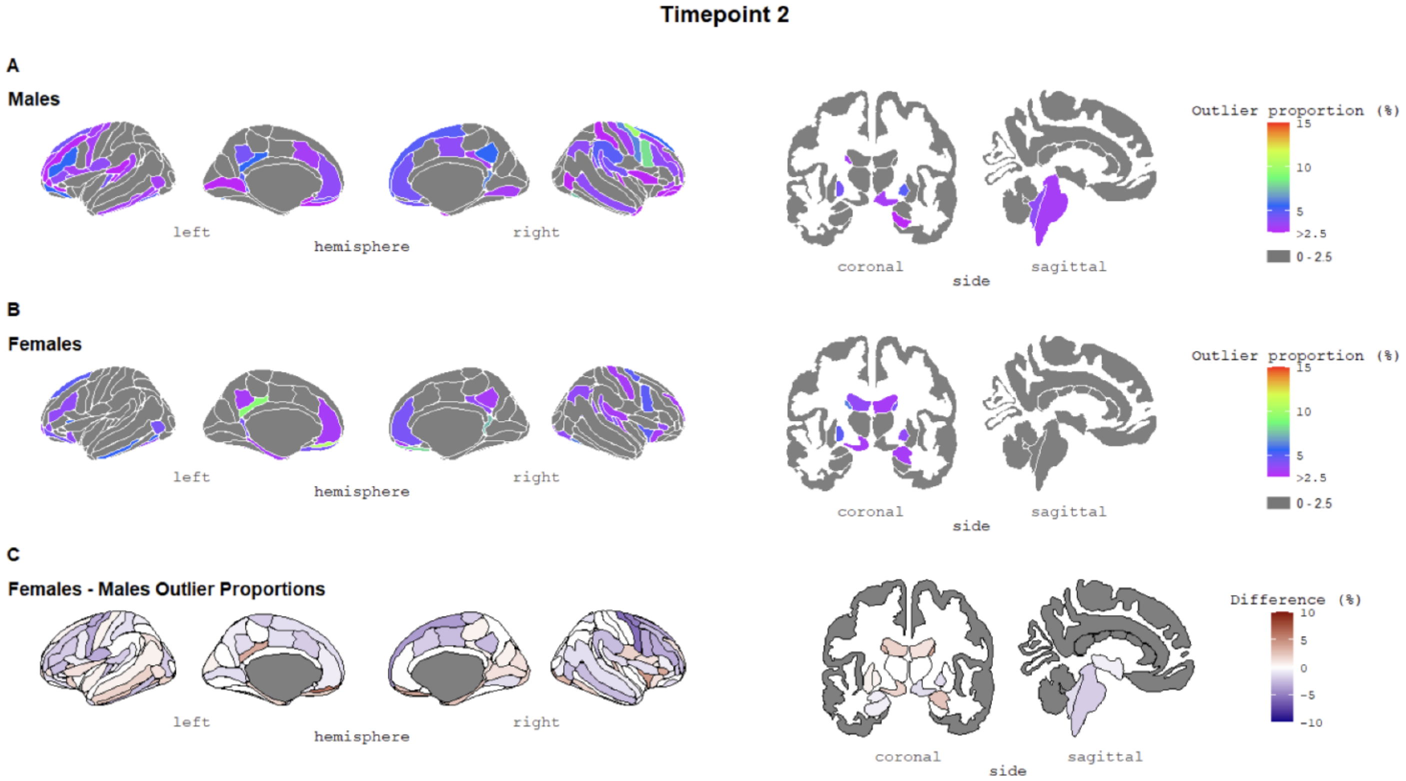


**SI Figure 4.** The percentage of outliers present within (A) females and (B) males at timepoint 2. The colour bar represents the outlier proportion (thresholding of z-scores). Regions in grey reflect areas where participants did not have any outliers (≤ 2.5 %). (C) The difference in outlier proportions between females and males. The colour gradient indicates the direction and magnitude of the difference: dark blue represents regions where males have more outliers than females, white indicates no difference between sexes, and dark red represents regions where females have more outliers than males. Color scale truncated at 15% to enhance visibility of regional variation. Same data visualized with full 0–100% scale for reference in Figure 2 of the main manuscript.

**Section 3. Model Regression Outputs**

**Note:** The reference category for Sex is females. The reference category for *APOE4* is non-carriers.

**SI Table 2.** Regression outputs for model 2 of the main manuscript

$$tOC = \beta_0 + \beta_1 Sex + \beta_2 Age + \beta_3 APOE4 + \beta_4 Sex \times Age \times APOE4 + u + \epsilon$$

| Regression term | Estimate | SE | Z-value | p-value |
|---|---|---|---|---|
| Intercept | 0.81 | 0.10 | 8.02 | $1.09 \times 10^{-15}$ |
| Age | -0.01 | 0.05 | -0.14 | 0.89 |
| Sex | 0.11 | 0.14 | 0.82 | 0.41 |
| APOE4 | -0.04 | 0.18 | -0.21 | 0.84 |
| Age*Sex | -0.17 | 0.06 | -2.59 | 0.01 |
| Age*APOE4 | -0.04 | 0.09 | -0.47 | 0.64 |
| Sex*APOE4 | 0.17 | 0.24 | 0.72 | 0.47 |
| Age*Sex*APOE4 | 0.13 | 0.12 | 1.14 | 0.26 |

**SI Table 3.** Regression output for the two-way interaction on tOC

$$tOC = \beta_0 + \beta_1 Age + \beta_2 APOE4 + \beta_3 Age \times APOE4 + u + \epsilon$$

| Regression term | Estimate | SE | Z-value | p-value |
|---|---|---|---|---|
| Intercept | 0.88 | 0.07 | 12.26 | $< 2 \times 10^{-16}$ |
| Age | -0.09 | 0.03 | -2.89 | 0.004 |
| APOE4 | 0.05 | 0.12 | 0.44 | 0.66 |
| Age*APOE4 | 0.03 | 0.06 | 0.44 | 0.66 |

**SI Table 4.** Regression output for the two-way interaction on tOC

$$tOC = \beta_0 + \beta_1 Sex + \beta_2 APOE4 + \beta_3 Sex \times APOE4 + u + \epsilon$$

| Regression term | Estimate | SE | Z-value | p-value |
|---|---|---|---|---|
| Intercept | 0.81 | 0.10 | 8.02 | $1.05 \times 10^{-15}$ |
| Sex | 0.13 | 0.14 | 0.96 | 0.34 |
| APOE4 | -0.03 | 0.17 | -0.17 | 0.87 |
| Sex*APOE4 | 0.16 | 0.24 | 0.67 | 0.50 |

**SI Table 5.** Regression outputs for model 2 of the main manuscript

$$tOC = \beta_0 + \beta_1 Sex + \beta_2 Age + \beta_3 Amyloid\ Centiloid + \beta_4 Sex \times Age \times Amyloid + u + \epsilon$$

| Regression term | Estimate | SE | Z-value | p-value |
|---|---|---|---|---|
| Intercept | 0.81 | 0.08 | 9.63 | $< 2 \times 10^{-16}$ |
| Age | -0.02 | 0.04 | -0.57 | 0.57 |
| Sex | 0.17 | 0.11 | 1.48 | 0.14 |
| Amyloid | 0.04 | 0.08 | 0.51 | 0.61 |
| Age*Sex | -0.13 | 0.05 | -2.33 | 0.02 |
| Age*Amyloid | 0.004 | 0.04 | 0.11 | 0.91 |
| Sex*Amyloid | 0.09 | 0.11 | 0.87 | 0.39 |
| Age*Sex* Amyloid | -0.02 | 0.05 | -0.36 | 0.72 |

**SI Table 6.** Regression output for the two-way interaction on tOC

$$tOC = \beta_0 + \beta_1 Age + \beta_2 Amyloid\ Centiloid + \beta_3\ Age \times Amyloid + u + \epsilon$$

| Regression term | Estimate | SE | Z-value | p-value |
|---|---|---|---|---|
| Intercept | 0.90 | 0.06 | 15.23 | $< 2 \times 10^{-16}$ |
| Age | -0.10 | 0.03 | -3.47 | 0.0005 |
| Amyloid | 0.10 | 0.05 | 1.83 | 0.07 |
| Age*Amyloid | -0.01 | 0.03 | -0.50 | 0.62 |

**SI Table 7.** Regression output for the two-way interaction on tOC

$$tOC = \beta_0 + \beta_1 Sex + \beta_2 Amyloid\ Centiloid + \beta_3\ Sex \times Amyloid + u + \epsilon$$

| Regression term | Estimate | SE | Z-value | p-value |
|---|---|---|---|---|
| Intercept | 0.81 | 0.08 | 9.70 | $< 2 \times 10^{-16}$ |
| Sex | 0.19 | 0.11 | 1.66 | 0.10 |
| Amyloid | 0.03 | 0.07 | 0.47 | 0.64 |
| Sex*Amyloid | 0.06 | 0.10 | 0.57 | 0.57 |

**SI Table 8.** Regression outputs for model 3 of the main manuscript (full sample)

$$PACC = \beta_0 + \beta_1 Sex + \beta_2 Amyloid\ Centiloid + \beta_3 tOC + \beta_4 Age + \beta_5 Sex \times Amyloid\ Centiloid \times tOC + u + \epsilon$$

| Regression term | Estimate | SE | Df | T-value | p-value |
|---|---|---|---|---|---|
| Intercept | 0.22 | 0.05 | 360.55 | 4.84 | $< 1.98 \times 10^{-6}$ |
| tOC | -0.05 | 0.04 | 671.33 | -1.41 | 0.16 |
| Amyloid | -0.08 | 0.04 | 540.04 | -2.09 | 0.04 |
| Sex | -0.33 | 0.06 | 362.69 | -5.20 | $3.42 \times 10^{-7}$ |
| Age | -0.01 | 0.01 | 343.65 | -0.81 | 0.42 |
| tOC*Amyloid | -0.10 | 0.04 | 611.38 | -2.71 | 0.007 |
| tOC*Sex | 0.06 | 0.05 | 652.66 | 1.20 | 0.23 |
| Amyloid*Sex | 0.07 | 0.06 | 537.12 | 1.25 | 0.21 |
| tOC*Amyloid*Sex | 0.11 | 0.04 | 611.88 | 2.52 | 0.01 |

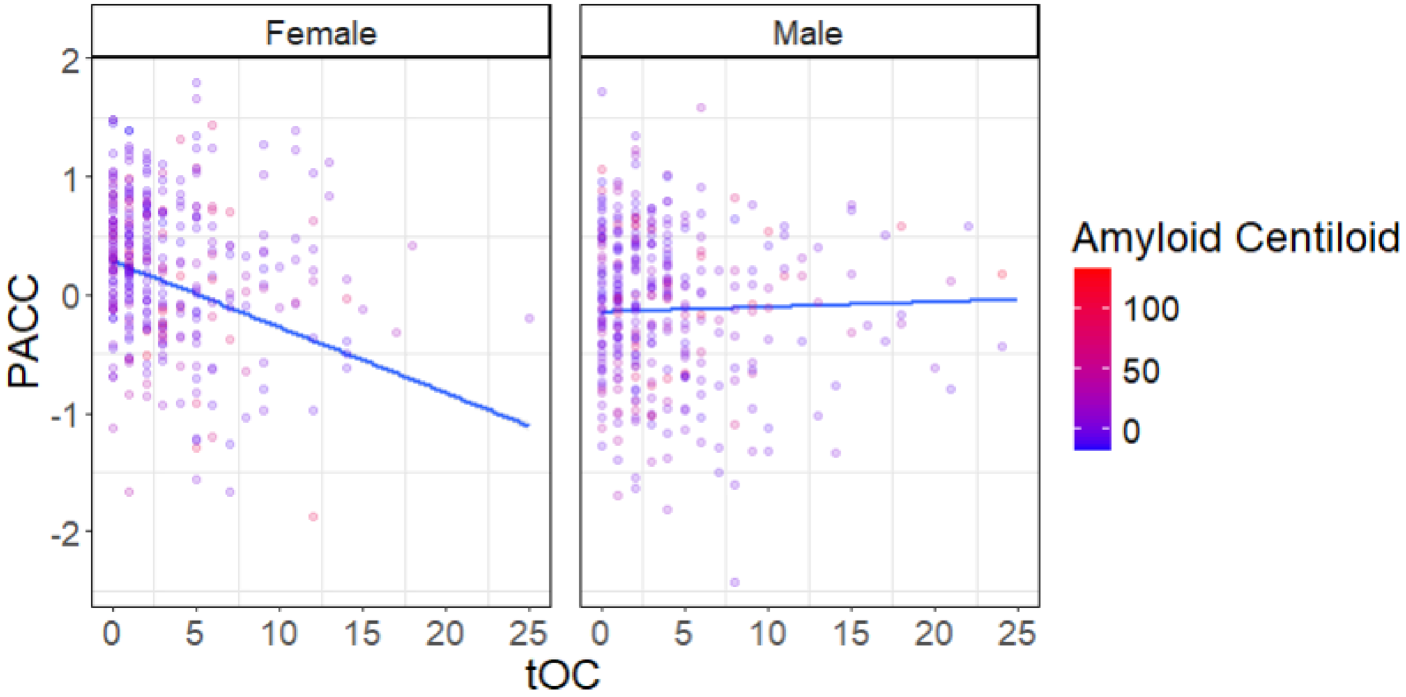


**SI Figure 5.** Fitted linear relationships between tOC and PACC scores in females and males, with their corresponding 95% confidence intervals (model 3). Data points on the plots represent raw data from the dataset and are colour-coded based on amyloid centiloid. Abbreviations: PACC = Preclinical Alzheimer Cognitive Composite; tOC = total outlier count.

**SI Table 9.** Regression output for the two-way interaction on PACC (full sample)

$$PACC = \beta_0 + \beta_1 Amyloid\ Centiloid + \beta_2 tOC + \beta_3 Age + \beta_4 Amyloid\ Centiloid \times tOC + u + \epsilon$$

| Regression term | Estimate | SE | Df | T-value | p-value |
|---|---|---|---|---|---|
| Intercept | 0.05 | 0.03 | 363.39 | 1.58 | 0.11 |
| tOC | -0.02 | 0.02 | 601.01 | -0.79 | 0.43 |
| Amyloid | -0.04 | 0.03 | 508.92 | -1.27 | 0.20 |
| Age | -0.01 | 0.01 | 342.41 | -0.95 | 0.35 |
| tOC*Amyloid | -0.02 | 0.02 | 615.54 | -1.05 | 0.30 |

**SI Table 10.** Regression output for the two-way interaction on PACC (full sample)

$$PACC = \beta_0 + \beta_1 Sex + \beta_2 tOC + \beta_3 Age + \beta_4 Sex \times tOC + u + \epsilon$$

| Regression term | Estimate | SE | Df | T-value | p-value |
|---|---|---|---|---|---|
| Intercept | 0.21 | 0.05 | 368.84 | 4.65 | $4.60 \times 10^{-6}$ |
| tOC | -0.06 | 0.04 | 687.89 | -1.42 | 0.16 |
| Sex | -0.33 | 0.06 | 371.27 | -5.22 | $2.98 \times 10^{-7}$ |
| Age | -0.01 | 0.01 | 328.74 | -1.17 | 0.24 |
| tOC*Sex | 0.05 | 0.05 | 674.78 | 1.10 | 0.27 |

**SI Table 11.** Regression output for the two-way interaction on PACC (full sample)

$$PACC = \beta_0 + \beta_1 Sex + \beta_2 Amyloid\ Centiloid + \beta_3 Age + \beta_4 Sex \times Amyloid\ Centiloid + u + \epsilon$$

| Regression term | Estimate | SE | Df | T-value | p-value |
|---|---|---|---|---|---|
| Intercept | 0.22 | 0.05 | 367.67 | 4.87 | $1.64 \times 10^{-6}$ |
| Amyloid | -0.08 | 0.04 | 542.47 | -1.90 | 0.06 |
| Sex | -0.33 | 0.06 | 369.79 | -5.21 | $3.13 \times 10^{-7}$ |
| Age | -0.01 | 0.01 | 348.55 | -0.86 | 0.39 |
| Amyloid*Sex | 0.07 | 0.06 | 529.33 | 1.17 | 0.24 |

**SI Table 12.** Regression outputs for model 3 of the main manuscript (*APOE4* non-carriers)

$$PACC = \beta_0 + \beta_1 Sex + \beta_2 Amyloid\ Centiloid + \beta_3 tOC + \beta_4 Age + \beta_5 Sex \times Amyloid\ Centiloid \times tOC + u + \epsilon$$

| Regression term | Estimate | SE | Df | T-value | p-value |
|---|---|---|---|---|---|
| Intercept | 0.26 | 0.06 | 241.41 | 4.54 | $8.94 \times 10^{-6}$ |
| tOC | -0.01 | 0.05 | 450.13 | -0.24 | 0.81 |
| Amyloid | -0.05 | 0.06 | 435.42 | -0.80 | 0.43 |
| Sex | -0.45 | 0.08 | 242.74 | -5.62 | $5.21 \times 10^{-8}$ |
| Age | -0.01 | 0.02 | 232.51 | -0.84 | 0.40 |
| tOC* amyloid | -0.02 | 0.07 | 370.14 | -0.31 | 0.76 |
| tOC*Sex | 0.02 | 0.06 | 426.55 | 0.36 | 0.72 |
| amyloid*Sex | -0.04 | 0.09 | 394.30 | -0.49 | 0.63 |
| tOC*Amyloid*Sex | 0.04 | 0.09 | 352.92 | 0.41 | 0.68 |

**SI Table 13.** Regression output for the two-way interaction on PACC (*APOE4* non-carriers)

$$PACC = \beta_0 + \beta_1 Amyloid\ Centiloid + \beta_2 tOC + \beta_3 Age + \beta_4 Amyloid\ Centiloid \times tOC + u + \epsilon$$

| Regression term | Estimate | SE | Df | T-value | p-value |
|---|---|---|---|---|---|
| Intercept | 0.03 | 0.04 | 244.60 | 0.76 | 0.45 |
| tOC | 0.002 | 0.03 | 375.02 | 0.06 | 0.95 |
| Amyloid | -0.06 | 0.05 | 393.99 | 1.38 | 0.17 |
| Age | -0.01 | 0.01 | 232.00 | -0.79 | 0.43 |
| tOC*Amyloid | -0.01 | 0.04 | 324.54 | -0.20 | 0.84 |

**SI Table 14.** Regression output for the two-way interaction on PACC (*APOE4* non-carriers)

$$PACC = \beta_0 + \beta_1 Sex + \beta_2 tOC + \beta_3 Age + \beta_4 Sex \times tOC + u + \epsilon$$

| Regression term | Estimate | SE | Df | T-value | p-value |
|---|---|---|---|---|---|
| Intercept | 0.27 | 0.06 | 236.32 | 4.83 | $2.47 \times 10^{-6}$ |
| tOC | -0.005 | 0.04 | 449.89 | -0.10 | 0.92 |
| Sex | -0.44 | 0.08 | 237.79 | -5.62 | $5.22 \times 10^{-8}$ |
| Age | -0.02 | 0.01 | 221.35 | -1.17 | 0.24 |
| tOC*Sex | 0.01 | 0.05 | 425.88 | 0.16 | 0.87 |

**SI Table 15.** Regression output for the two-way interaction on PACC (*APOE4* non-carriers)

$$PACC = \beta_0 + \beta_1 Sex + \beta_2 Amyloid\ Centiloid + \beta_3 Age + \beta_4 Sex \times Amyloid\ Centiloid + u + \epsilon$$

| Regression term | Estimate | SE | Df | T-value | p-value |
|---|---|---|---|---|---|
| Intercept | 0.26 | 0.06 | 247.75 | 4.59 | $7.07 \times 10^{-6}$ |
| Amyloid | -0.04 | 0.06 | 432.90 | -0.73 | 0.46 |
| Sex | -0.45 | 0.08 | 249.29 | -5.65 | $4.32 \times 10^{-8}$ |
| Age | -0.01 | 0.01 | 239.25 | -0.93 | 0.35 |
| Amyloid*Sex | -0.05 | 0.09 | 398.77 | -0.57 | 0.57 |

**SI Table 16.** Regression outputs for model 3 of the main manuscript (*APOE4* carriers)

$$PACC = \beta_0 + \beta_1 Sex + \beta_2 Amyloid\ Centiloid + \beta_3 tOC + \beta_4 Age + \beta_5 Sex \times Amyloid\ Centiloid \times tOC + u + \epsilon$$

| Regression term | Estimate | SE | Df | T-value | p-value |
|---|---|---|---|---|---|
| Intercept | 0.15 | 0.09 | 123.46 | 1.69 | 0.09 |
| tOC | -0.13 | 0.09 | 180.86 | -1.40 | 0.16 |
| Amyloid | -0.07 | 0.06 | 148.04 | -1.18 | 0.24 |
| Sex | -0.15 | 0.12 | 125.38 | -1.25 | 0.21 |
| Age | -0.005 | 0.02 | 105.48 | -0.21 | 0.84 |
| tOC*Amyloid | -0.11 | 0.05 | 198.76 | -2.05 | 0.04 |
| tOC*Sex | 0.11 | 0.11 | 199.38 | 1.08 | 0.28 |
| Amyloid*Sex | 0.08 | 0.09 | 162.34 | 0.98 | 0.33 |
| tOC*Amyloid*Sex | 0.11 | 0.06 | 196.72 | 1.87 | 0.06 |

**SI Table 17.** Regression output for the two-way interaction on PACC (*APOE4* carriers)

$$PACC = \beta_0 + \beta_1 Amyloid\ Centiloid + \beta_2 tOC + \beta_3 Age + \beta_4 Amyloid\ Centiloid \times tOC + u + \epsilon$$

| Regression term | Estimate | SE | Df | T-value | p-value |
|---|---|---|---|---|---|
| Intercept | 0.07 | 0.06 | 125.71 | 1.20 | 0.23 |
| tOC | -0.05 | 0.05 | 203.35 | -1.15 | 0.25 |
| Amyloid | -0.02 | 0.04 | 145.81 | -0.48 | 0.63 |
| Age | -0.006 | 0.03 | 106.62 | -0.23 | 0.82 |
| tOC*Amyloid | -0.02 | 0.03 | 200.79 | -0.61 | 0.54 |

**SI Table 18.** Regression output for the two-way interaction on PACC (*APOE4* carriers)

$$PACC = \beta_0 + \beta_1 Sex + \beta_2 tOC + \beta_3 Age + \beta_4 Sex \times tOC + u + \epsilon$$

| Regression term | Estimate | SE | Df | T-value | p-value |
|---|---|---|---|---|---|
| Intercept | 0.10 | 0.08 | 124.10 | 1.24 | 0.22 |
| tOC | -0.21 | 0.08 | 188.21 | -2.56 | 0.01 |
| Sex | -0.10 | 0.11 | 124.13 | -0.84 | 0.40 |
| Age | -0.01 | 0.02 | 95.84 | -0.47 | 0.64 |
| tOC*Sex | 0.20 | 0.10 | 202.87 | 2.08 | 0.04 |

**SI Table 19.** Regression output for the two-way interaction on PACC (*APOE4* carriers)

$$PACC = \beta_0 + \beta_1 Sex + \beta_2 Amyloid\ Centiloid + \beta_3 Age + \beta_4 Sex \times Amyloid\ Centiloid + u + \epsilon$$

| Regression term | Estimate | SE | Df | T-value | p-value |
|---|---|---|---|---|---|
| Intercept | 0.16 | 0.09 | 124.87 | 1.82 | 0.07 |
| Amyloid | -0.07 | 0.06 | 149.21 | -1.29 | 0.20 |
| Sex | -0.16 | 0.12 | 127.07 | -1.36 | 0.18 |
| Age | -0.004 | 0.03 | 107.49 | -0.16 | 0.87 |
| Amyloid*Sex | 0.09 | 0.08 | 157.63 | 1.12 | 0.27 |

**SI Table 20.** Regression outputs for model 5 of the main manuscript

$$tOC\ Change = \beta_0 + \beta_1 Sex + \beta_2 Age\ Timepoint\ 1 + \beta_3 APOE4 + \beta_4 Sex \times Age\ Timepoint\ 1 \times APOE4 + \beta_5 Assessment\ Interval + \epsilon$$

| Regression Term | Estimate | SE | T-value | p-value |
|---|---|---|---|---|
| Intercept | -0.01 | 0.29 | -0.05 | 0.96 |
| Age TP1 | 0.08 | 0.28 | 0.29 | 0.77 |
| Sex | -0.93 | 0.41 | -2.30 | 0.02 |
| APOE4 | 0.008 | 0.53 | 0.02 | 0.99 |
| Assessment Interval | -0.24 | 0.17 | -1.37 | 0.17 |
| Age TP1*Sex | 2.10 | 0.41 | 5.14 | <0.05 |
| Age TP1*APOE4 | 0.34 | 0.56 | 0.61 | 0.54 |
| Sex*APOE4 | 0.83 | 0.75 | 1.11 | 0.27 |
| Age TP1*Sex*APOE4 | -0.57 | 0.76 | -0.75 | 0.46 |

Note: Change was measured using difference scores [Timepoint 2 values − Timepoint 1 values]. TP1 = Timepoint 1

**SI Table 21.** Regression outputs for model 5 of the main manuscript

$$PACC\ Change = \beta_0 + \beta_1 Sex + \beta_2 Age\ Timepoint\ 1 + \beta_3 APOE4 + \beta_4 Sex \times Age\ Timepoint\ 1 \times APOE4 + \beta_5 Assessment\ Interval + \epsilon$$

| Regression Term | Estimate | SE | T-value | p-value |
|---|---|---|---|---|
| Intercept | -0.05 | 0.04 | -1.34 | 0.18 |
| Age TP1 | -0.02 | 0.04 | -0.50 | 0.62 |
| Sex | 0.02 | 0.05 | 0.31 | 0.76 |
| APOE4 | 0.04 | 0.07 | 0.57 | 0.57 |
| Assessment Interval | 0.02 | 0.02 | 0.81 | 0.42 |
| Age TP1*Sex | 0.05 | 0.06 | 0.93 | 0.35 |
| Age TP1*APOE4 | 0.03 | 0.08 | 0.41 | 0.68 |
| Sex*APOE4 | -0.01 | 0.10 | -0.08 | 0.94 |
| Age TP1*Sex*APOE4 | -0.06 | 0.10 | -0.55 | 0.58 |

Note: Change was measured using difference scores [Timepoint 2 values − Timepoint 1 values]. TP1 = Timepoint 1.

**SI Table 22.** Regression outputs for model 5 of the main manuscript

$$tOC\ Change\ =\ \beta_0 +\ \beta_1 Sex + \beta_2 Age\ Timepoint\ 1 + \beta_3 Amyloid\ Centiloid +\ \ \beta_4 Sex\ \times\ Age\ Timepoint\ 1\ \times\ Amyloid\ Centiloid + \beta_5 Assessment\ Interval\ + \epsilon$$

| Regression term | Estimate | SE | T-value | p-value |
|---|---|---|---|---|
| Intercept | -0.06 | 0.29 | -0.19 | 0.85 |
| Age TP1 | 0.20 | 0.29 | 0.70 | 0.48 |
| Sex | -0.54 | 0.41 | -1.33 | 0.19 |
| Amyloid 1 | 0.54 | 0.60 | 0.91 | 0.37 |
| Amyloid 2 | -1.11 | 0.91 | -1.22 | 0.22 |
| Assessment Interval | -0.25 | 0.18 | -1.40 | 0.16 |
| Age TP1*Sex | 2.13 | 0.40 | 5.26 | $2.87 \times 10^{-7}$ |
| Age TP1*Amyloid 1 | -0.31 | 0.66 | -0.48 | 0.64 |
| Age TP1*Amyloid 2 | 0.50 | 1.03 | 0.49 | 0.62 |
| Sex*Amyloid 1 | -0.88 | 0.85 | -1.04 | 0.30 |
| Sex*Amyloid 2 | 0.62 | 1.50 | 0.41 | 0.68 |
| Age TP1*Sex*Amyloid 1 | -0.47 | 0.88 | -0.53 | 0.60 |
| Age TP1*Sex*Amyloid 2 | -0.22 | 1.60 | -1.39 | 0.17 |

Note: Change was measured using difference scores [Timepoint 2 values − Timepoint 1 values]. The reference group for Amyloid is 0 (participants with no amyloid at either timepoint). Amyloid 1 = amyloid present at both timepoints; Amyloid 2 = amyloid negative at timepoint 1 but positive at timepoint 2 (converters). TP1 = Timepoint 1.

**SI Table 23.** Regression outputs for model 5 of the main manuscript

$$PACC\ Change\ =\ \beta_0 +\ \beta_1 Sex + \beta_2 Age\ Timepoint\ 1 + \beta_3 Amyloid\ Centiloid +\ \ \beta_4 Sex\ \times\ Age\ Timepoint\ 1\ \times\ Amyloid\ Centiloid + \beta_5 Assessment\ Interval\ + \epsilon$$

| Regression term | Estimate | SE | T-value | p-value |
|---|---|---|---|---|
| Intercept | -0.05 | 0.04 | -1.25 | 0.21 |
| Age TP1 | -0.02 | 0.04 | -0.47 | 0.64 |
| Sex | 0.01 | 0.06 | 0.13 | 0.90 |
| Amyloid 1 | -0.02 | 0.08 | -0.31 | 0.76 |
| Amyloid 2 | 0.08 | 0.12 | 0.65 | 0.52 |
| Assessment Interval | 0.01 | 0.02 | 0.61 | 0.54 |
| Age TP1*Sex | 0.03 | 0.05 | 0.57 | 0.57 |
| Age TP1*Amyloid 1 | 0.01 | 0.09 | 0.15 | 0.88 |
| Age TP1*Amyloid 2 | 0.011 | 0.14 | 0.08 | 0.93 |
| Sex*Amyloid 1 | 0.13 | 0.11 | 1.18 | 0.24 |
| Sex*Amyloid 2 | -0.23 | 0.20 | -1.12 | 0.26 |
| Age TP1*Sex*Amyloid 1 | 0.04 | 0.12 | 0.37 | 0.71 |
| Age TP1*Sex*Amyloid 2 | 0.20 | 0.21 | 0.93 | 0.35 |

Note: Change was measured using difference scores [Timepoint 2 values − Timepoint 1 values]. The reference group for Amyloid is 0 (participants with no amyloid at either timepoint). Amyloid 1 = amyloid present at both timepoints; Amyloid 2 = amyloid negative at timepoint 1 but positive at timepoint 2 (converters). TP1 = Timepoint 1.

**Section 4. Alternative Longitudinal Analyses**

As an alternative measure of change in tOC/PACC, we reran models 4 and 5 using timepoint 2 values with timepoint 1 values regressed out (i.e., modelling changes in DV independent of its initial value; Gollwitzer et al., 2014). This approach can be useful when timepoint 1 tOC/PACC values vary substantially between participants, as the residualized change helps account for timepoint 1 differences (Vickers & Altman, 2001).

**Model 4.1:**

$$DV\ Change = \beta_0 + \beta_1 Sex + \beta_2 Age\ Timepoint\ 1 + \beta_3 Sex \times Age\ Timepoint\ 1 + \\ + \beta_4 Assessment\ Interval + \epsilon$$

**Model 5.1:**

$$DV\ Change = \beta_0 + \beta_1 Sex + \beta_2 Age\ Timepoint\ 1 + \beta_3 AD\ Risk + \\ \beta_4 Sex \times Age\ Timepoint\ 1 \times AD\ Risk + \beta_5 Assessment\ Interval + \epsilon$$

All other predictor variables were treated in the same way as in the primary analyses.

Our results were consistent with the primary findings. Model 4.1 revealed a significant interaction between age and sex on change on tOC ($\beta$ = 1.82, SE = 0.33, z = 5.50, p = <0.01), indicating that an older age was related to a greater change in tOC, with stronger effects in males compared to females (Supplementary Figure 2). When changes in PACC scores were used as the outcome, this model did not reveal any significant effects of age ($\beta$ = -0.02, SE = 0.03, t = -0.59, p = 0.56), sex ($\beta$ = -0.05, SE = 0.04, t = -1.20, p = 0.23), or their interaction ($\beta$ = 0.04, SE = 0.04, t = 1.01, p = 0.31).

Model 5.1 did not reveal a significant interaction between age, sex, and *APOE4* status on changes in tOC or PACC. Model 5.1 using amyloid values as the AD risk factor showed no significant effect of age, sex, amyloid values or their interactions on changes in tOC or PACC. Refer to Supplementary Table 25-28 for a detailed list of regression outputs from this model.

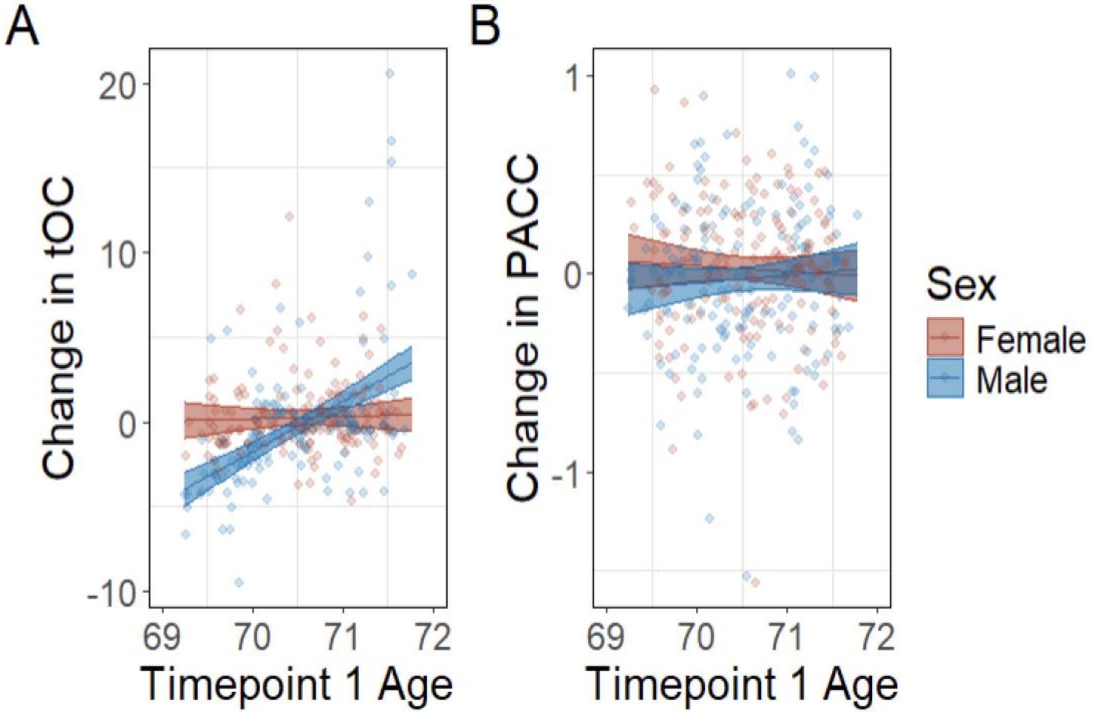


**SI Figure 6.** Fitted relationships between age at timepoint 1 and changes in tOC (A) or PACC (B), with corresponding 95% confidence intervals (model 4). Change was measured using timepoint 2 values with timepoint 1 values regressed out. Data points on the plots represent raw data from the dataset. Abbreviations: PACC = Preclinical Alzheimer Cognitive Composite; tOC = total outlier count.

**SI Table 24.** Regression outputs for model 5.1 of the main manuscript

$$tOC\ Change\ =\ \beta_0 +\ \beta_1 Sex + \beta_2 Age\ Timepoint\ 1 + \beta_3 AD\ Risk +\ \beta_4 Sex\ \times\ Age\ Timepoint\ 1\ \times\ APOE4\ +\ \beta_5 Assessment\ Interval\ + \epsilon$$

| **Regression Term** | **Estimate** | **SE** | **T-value** | **p-value** |
|---|---|---|---|---|
| Intercept | 0.31 | 0.28 | 1.12 | 0.26 |
| Age TP1 | -0.03 | 0.28 | -0.12 | 0.90 |
| Sex | -0.76 | 0.40 | -1.92 | 0.06 |
| APOE4 | -0.08 | 0.52 | -0.15 | 0.88 |
| Assessment Interval | -0.14 | 0.17 | -0.83 | 0.41 |
| Age TP1*Sex | 2.02 | 0.40 | 5.05 | $7.84 \times 10^{-7}$ |
| Age TP1*APOE4 | 0.43 | 0.55 | 0.79 | 0.43 |
| Sex*APOE4 | 0.90 | 0.73 | 1.23 | 0.22 |
| Age TP1*sex*APOE4 | -0.50 | 0.74 | -0.68 | 0.50 |

Note: Change was measured using timepoint 2 values with timepoint 1 values regressed out. TP1 = Timepoint 1.

**SI Table 25**. Regression outputs for model 5.1 of the main manuscript

$$PACC\ Change\ = \beta_0 + \beta_1 Sex + \beta_2 Age\ Timepoint\ 1 + \beta_3 AD\ Risk + \beta_4 Sex \times Age\ Timepoint\ 1 \times APOE4 + \beta_5 Assessment\ Interval\ + \epsilon$$

| Regression Term | Estimate | SE | T-value | p-value |
|---|---|---|---|---|
| Intercept | 0.03 | 0.04 | 0.76 | 0.45 |
| Age TP1 | -0.02 | 0.04 | -0.48 | 0.64 |
| Sex | -0.06 | 0.05 | -1.22 | 0.22 |
| APOE4 | -0.004 | 0.07 | -0.06 | 0.95 |
| Assessment Interval | 0.02 | 0.02 | 0.77 | 0.44 |
| Age TP1*Sex | 0.05 | 0.05 | 0.95 | 0.34 |
| Age TP1*APOE4 | -0.005 | 0.07 | -0.07 | 0.94 |
| Sex*APOE4 | 0.05 | 0.10 | 0.47 | 0.64 |
| Age TP1*sex*APOE4 | -0.02 | 0.10 | -0.17 | 0.87 |

Note: Change was measured using timepoint 2 values with timepoint 1 values regressed out. TP1 = Timepoint 1.

**SI Table 26.** Regression outputs for model 5.1 of the main manuscript

$$tOC\ Change\ = \beta_0 + \beta_1 Sex + \beta_2 Age\ Timepoint\ 1 + \beta_3 AD\ Risk + \beta_4 Sex \times Age\ Timepoint\ 1 \times Amyloid + \beta_5 Assessment\ Interval\ + \epsilon$$

| Regression term | Estimate | SE | T-value | p-value |
|---|---|---|---|---|
| Intercept | 0.23 | 0.28 | 0.80 | 0.43 |
| Age TP1 | 0.15 | 0.28 | 0.54 | 0.59 |
| Sex | -0.36 | 0.40 | -0.89 | 0.37 |
| Amyloid 1 | 0.50 | 0.59 | 0.85 | 0.40 |
| Amyloid 2 | -0.78 | 0.89 | -0.87 | 0.38 |
| Assessment Interval | -0.16 | 0.17 | -0.93 | 0.35 |
| Age TP1*Sex | 2.06 | 0.40 | 5.20 | $3.77 \times 10^{-7}$ |
| Age TP1*Amyloid 1 | -0.39 | 0.65 | -0.60 | 0.55 |
| Age TP1*Amyloid 2 | -0.05 | 1.00 | -0.05 | 0.96 |
| Sex*Amyloid 1 | -0.66 | 0.83 | -0.79 | 0.43 |
| Sex*Amyloid 2 | 0.13 | 1.47 | 0.09 | 0.93 |
| Age TP1*Sex*Amyloid 1 | -0.48 | 0.86 | -0.55 | 0.58 |
| Age TP1*Sex*Amyloid 2 | -1.84 | 1.56 | -1.18 | 0.24 |

Note: Change was measured using timepoint 2 values with timepoint 1 values regressed out. The reference group for Amyloid is 0 (participants with no amyloid at either timepoint). Amyloid 1 = amyloid present at both timepoints; Amyloid 2 = amyloid negative at timepoint 1 but positive at timepoint 2 (converters). TP1 = Timepoint 1.

**SI Table 27.** Regression outputs for model 5.1 of the main manuscript

$$PACC\ Change\ =\ \beta_0 +\ \beta_1 Sex + \beta_2 Age\ Timepoint\ 1 + \beta_3 AD\ Risk +\ \beta_4 Sex\ \times\ Age\ Timepoint\ 1\ \times\ Amyloid + \beta_5 Assessment\ Interval\ + \epsilon$$

| Regression term | Estimate | SE | T-value | p-value |
|---|---|---|---|---|
| Intercept | 0.03 | 0.04 | 0.85 | 0.40 |
| Age TP1 | -0.01 | 0.04 | -0.29 | 0.78 |
| Sex | -0.07 | 0.05 | -1.26 | 0.21 |
| Amyloid 1 | -0.03 | 0.08 | -0.43 | 0.67 |
| Amyloid 2 | -0.004 | 0.12 | -0.04 | 0.97 |
| Assessment Interval | 0.01 | 0.02 | 0.66 | 0.51 |
| Age TP1*Sex | 0.03 | 0.05 | 0.53 | 0.60 |
| Age TP1*Amyloid 1 | -0.04 | 0.08 | -0.53 | 0.60 |
| Age TP1*Amyloid 2 | 0.04 | 0.13 | 0.28 | 0.78 |
| Sex*Amyloid 1 | 0.12 | 0.11 | 1.10 | 0.27 |
| Sex*Amyloid 2 | -0.15 | 0.19 | -0.77 | 0.44 |
| Age TP1*Sex*Amyloid 1 | 0.07 | 0.11 | 0.60 | 0.55 |
| Age TP1*Sex*Amyloid 2 | 0.18 | 0.20 | 0.90 | 0.37 |

Note: Change was measured using timepoint 2 values with timepoint 1 values regressed out. The reference group for Amyloid is 0 (participants with no amyloid at either timepoint). Amyloid 1 = amyloid present at both timepoints; Amyloid 2 = amyloid negative at timepoint 1 but positive at timepoint 2 (converters). TP1 = Timepoint 1.

**SI Section 5. Matched Longitudinal Analyses (Models 1-3)**

Of the participants used in the primary analyses, 41 were present at timepoint 1 but not at timepoint 2. This group did not exhibit substantially higher tOC values (mean = 4.2, SD = 4.19; range = 0-18; median = 4). While two participants had relatively high counts (17 and 18), the remaining values fell within a range of 0 to 11.

Supplementary analyses across all models using a matched longitudinal sample of participants with data available at both timepoints were consistent with the primary analyses (SI Tables 28-34).

**SI Table 28.** Regression output for model 1
$tOC = \beta_0 + \beta_1 Sex + \beta_2 Age + \beta_3 Sex \times Age + u + \epsilon$

| Regression term | Estimate | SE | Z-value | p-value |
|---|---|---|---|---|
| Intercept | 0.76 | 0.09 | 8.18 | $2.89 \times 10^{-16}$ |
| Age | -0.02 | 0.04 | -0.42 | 0.67 |
| Sex | 0.21 | 0.13 | 1.68 | 0.09 |
| Age*Sex | -0.13 | 0.06 | -2.33 | 0.02 |

**SI Table 29.** Regression output for model 1
$PACC = \beta_0 + \beta_1 Sex + \beta_2 Age + \beta_3 Sex \times Age + u + \epsilon$

| Regression term | Estimate | SE | Df | T-value | p-value |
|---|---|---|---|---|---|
| Intercept | 0.25 | 0.05 | 307.02 | 5.30 | $2.23 \times 10^{-7}$ |
| Age | -0.03 | 0.02 | 307.02 | -1.46 | 0.15 |
| Sex | -0.32 | 0.07 | 325.47 | 4.84 | $2.03 \times 10^{-6}$ |
| Age*Sex | 0.01 | 0.03 | 326.49 | 0.57 | 0.57 |

**SI Table 30.** Regression outputs for model 2 of the main manuscript
$tOC = \beta_0 + \beta_1 Sex + \beta_2 Age + \beta_3 APOE4 + \beta_4 Sex \times Age \times APOE4 + u + \epsilon$

| Regression term | Estimate | SE | Z-value | p-value |
|---|---|---|---|---|
| Intercept | 0.78 | 0.11 | 7.18 | $7.25 \times 10^{-13}$ |
| Age | -0.01 | 0.05 | -0.21 | 0.84 |
| Sex | 0.16 | 0.15 | 1.02 | 0.31 |
| APOE4 | -0.09 | 0.20 | -0.44 | 0.66 |
| Age*Sex | -0.16 | 0.07 | -2.39 | 0.02 |
| Age*APOE4 | -0.03 | 0.09 | -0.30 | 0.76 |
| Sex*APOE4 | 0.16 | 0.28 | 0.57 | 0.57 |
| Age*Sex*APOE4 | 0.11 | 0.12 | 0.87 | 0.39 |

**SI Table 31.** Regression outputs for model 2 of the main manuscript

$$tOC = \beta_0 + \beta_1 Sex + \beta_2 Age + \beta_3 Amyloid\ Centiloid + \beta_4 Sex \times Age \times Amyloid + u + \epsilon$$

| Regression term | Estimate | SE | Z-value | p-value |
|---|---|---|---|---|
| Intercept | 0.77 | 0.09 | 8.28 | $< 2 \times 10^{-16}$ |
| Age | -0.02 | 0.04 | -0.58 | 0.56 |
| Sex | 0.21 | 0.13 | 1.65 | 0.10 |
| Amyloid | 0.06 | 0.09 | 0.67 | 0.50 |
| Age*Sex | -0.13 | 0.06 | -2.27 | 0.02 |
| Age*Amyloid | -0.004 | 0.04 | -0.09 | 0.93 |
| Sex*Amyloid | 0.09 | 0.12 | 0.69 | 0.49 |
| Age*Sex* Amyloid | -0.01 | 0.05 | -0.16 | 0.87 |

**SI Table 32.** Regression outputs for model 3 of the main manuscript (full sample)

$$PACC = \beta_0 + \beta_1 Sex + \beta_2 Amyloid\ Centiloid + \beta_3 tOC + \beta_4 Age + \beta_5 Sex \times Amyloid\ Centiloid \times tOC + u + \epsilon$$

| Regression term | Estimate | SE | Df | T-value | p-value |
|---|---|---|---|---|---|
| Intercept | 0.25 | 0.05 | 300.24 | 5.36 | $1.66 \times 10^{-7}$ |
| tOC | -0.05 | 0.04 | 599.80 | -1.36 | 0.18 |
| Amyloid | -0.05 | 0.04 | 468.84 | -1.33 | 0.18 |
| Sex | -0.32 | 0.07 | 300.33 | -4.87 | $1.79 \times 10^{-6}$ |
| Age | -0.02 | 0.01 | 345.26 | -1.24 | 0.22 |
| tOC*Amyloid | -0.11 | 0.04 | 561.60 | -2.83 | 0.005 |
| tOC*Sex | 0.06 | 0.05 | 594.66 | 1.34 | 0.18 |
| Amyloid*Sex | 0.05 | 0.06 | 463.94 | 0.82 | 0.41 |
| tOC*Amyloid*Sex | 0.11 | 0.04 | 564.44 | 2.46 | 0.01 |

**SI Table 33.** Regression outputs for model 3 of the main manuscript (*APOE4* non-carriers)

$$PACC = \beta_0 + \beta_1 Sex + \beta_2 Amyloid\ Centiloid + \beta_3 tOC + \beta_4 Age + \beta_5 Sex \times Amyloid\ Centiloid \times tOC + u + \epsilon$$

| Regression term | Estimate | SE | Df | T-value | p-value |
|---|---|---|---|---|---|
| Intercept | 0.29 | 0.06 | 210.34 | 5.28 | $3.26 \times 10^{-7}$ |
| tOC | -0.01 | 0.05 | 416.97 | -0.25 | 0.80 |
| Amyloid | -0.03 | 0.06 | 387.99 | -0.55 | 0.58 |
| Sex | -0.42 | 0.08 | 211.76 | -5.11 | $7.13 \times 10^{-7}$ |
| Age | -0.02 | 0.02 | 235.52 | -1.14 | 0.26 |
| tOC*Amyloid | -0.03 | 0.07 | 357.40 | -0.43 | 0.67 |
| tOC*Sex | 0.02 | 0.06 | 405.92 | 0.36 | 0.72 |
| amyloid*Sex | -0.09 | 0.09 | 344.78 | -1.03 | 0.30 |
| tOC*Amyloid*Sex | 0.02 | 0.09 | 331.06 | 0.23 | 0.82 |

**SI Table 34.** Regression outputs for model 3 of the main manuscript (*APOE4* carriers)

$$PACC = \beta_0 + \beta_1 Sex + \beta_2 Amyloid\ Centiloid + \beta_3 tOC + \beta_4 Age + \beta_5 Sex \times Amyloid\ Centiloid \times tOC + u + \epsilon$$

| Regression term | Estimate | SE | Df | T-value | p-value |
|---|---|---|---|---|---|
| Intercept | 0.10 | 0.10 | 83.09 | 1.03 | 0.31 |
| tOC | -0.17 | 0.10 | 135.32 | -1.63 | 0.11 |
| Amyloid | 0.006 | 0.07 | 108.55 | 0.08 | 0.94 |
| Sex | -0.19 | 0.14 | 84.75 | -1.33 | 0.19 |
| Age | -0.02 | 0.03 | 103.32 | -0.75 | 0.46 |
| tOC*Amyloid | -0.11 | 0.06 | 161.16 | -2.02 | 0.04 |
| tOC*Sex | 0.19 | 0.12 | 155.39 | 1.60 | 0.11 |
| Amyloid*Sex | 0.09 | 0.10 | 119.76 | 0.96 | 0.34 |
| tOC*Amyloid*Sex | 0.10 | 0.06 | 160.21 | 1.52 | 0.13 |
